# Predicting Subjective Cognitive Decline on Future BRFSS Survey Years: An Open Multi-Language Machine Learning Benchmark

**DOI:** 10.64898/2026.08.18.26360709

**Authors:** Trang T Nguyen, Truong D Nguyen

## Abstract

**Background and Objectives:** Subjective cognitive decline (SCD), self-reported worsening confusion or memory over the past year, is a common early marker of cognitive concern with relevance for Alzheimer’s disease prevention and population health. Population-based machine learning benchmarks that respect temporal drift in public health surveillance remain limited. We developed a reusable multi-language prediction and interpretability framework for SCD using Behavioral Risk Factor Surveillance System (BRFSS) Cognitive Decline data.

**Methods:** We analyzed pooled national (n = 298,944) and New York (n = 30,366) cohorts with chronological train (2015-2019), validation (national: 2020-2022; New York: 2020-2021), and locked test (2023- 2024) splits. Nested LASSO identified stable predictors. Sixteen machine learning algorithms were compared under year-grouped cross-validation with SMOTE restricted to training folds. Four end-to-end Python/R pipelines (single-model or soft-voting) used validation-only isotonic calibration and Youden thresholding. Primary reporting pipelines were prespecified before test unlock (national: R tidymodels single-model; New York: Python single-model); algorithms within each pipeline were chosen by validation ROC-AUC. Post-hoc GLMs (national unweighted; New York design-weighted) and two training-only knowledge-graph layers supported interpretability.

**Results:** Locked-test discrimination was consistent across implementations (ROC-AUC approximately 0.76-0.77). Prespecified pipelines achieved test ROC-AUC 0.770 (95% CI 0.767- 0.773) nationally (R gradient boosting) and 0.762 (95% CI 0.746-0.777) in New York (Python AdaBoost). Soft-voting pipelines performed similarly (national 0.770; New York 0.757) and were treated as sensitivity benchmarks. Predicted probabilities were reasonably calibrated (Brier 0.118 nationally; 0.112 in New York), and higher scores among SCD-positive respondents persisted across survey years. Difficulty deciding, mental health, and functional health items ranked highest across permutation importance, SHAP, and GLMs. Respondents who reported no difficulty deciding (DECIDE = 2) had substantially lower odds of SCD than those who reported difficulty (DECIDE = 1; aOR approximately 0.13; FDR < 0.05). Training-only knowledge graphs likewise placed difficulty deciding nearest to SCD in both cohorts.

**Conclusions:** A temporally locked, multi-pipeline BRFSS benchmark yields stable future-year SCD risk ranking, usable calibrated probability scores that remain separated by SCD status across survey years, and convergent interpretability signals. The open implementation supports reproducible surveillance-oriented machine learning for cognitive health.

**Highlights:**

- Chronological BRFSS splits (2015-2024) with a locked 2023-2024 SCD test set.
- Python/R pipelines yield consistent locked-test ROC-AUC of about 0.76-0.77.
- Primary pipelines are prespecified; classifiers are chosen on validation only.
- Difficulty deciding is the top signal across ML importance, SHAP, GLM, and graphs.
- Open multi-language SCD benchmark with calibration and knowledge-graph maps.

## 1. Introduction

Subjective cognitive decline (SCD), characterized by self-perceived deterioration in memory or cognitive capacity [1], affects 11.2% of adults aged 45 years or older [2]. SCD is an early dementia indicator: individuals with SCD have approximately twofold higher five-year dementia risk than those without SCD [3]. In preclinical Alzheimer’s disease (AD), SCD has been linked to AD-related biomarkers [4] and proposed as a clinical manifestation of transitional AD Stage 2 [5]. SCD is also associated with functional limitations and poorer quality of life [6], motivating earlier identification. Because SCD is nonspecific and may reflect aging, psychological factors, or medication effects as well as neurodegeneration, accurate risk ranking remains challenging.

While SCD is primarily based on self-reported cognitive concerns, instruments such as the Mini-Mental State Examination (MMSE), Montreal Cognitive Assessment (MoCA), and Mini-Cog are commonly used to evaluate objective cognition [7]. Because individuals with SCD typically perform normally on objective testing [8], brief screens often have limited sensitivity for earliest decline. Prior machine-learning SCD studies have used multimodal MRI [9] or demographic, behavioral, and psychological variables [7], but many relied on smaller clinical cohorts with limited generalizability.

Although previous studies have demonstrated the potential of machine learning for SCD risk prediction [10], classification of SCD due to AD using neuroimaging biomarkers [11], and prediction of progression from SCD to mild cognitive impairment (MCI) or AD [12], population-based models developed using large-scale public health surveillance data remain limited. Furthermore, to our knowledge, previous machine learning studies of SCD have not reported a fixed year-based evaluation on future survey cycles together with an exportable multi-language software workflow. Therefore, we developed and evaluated machine learning models for SCD risk prediction using the Behavioral Risk Factor Surveillance System (BRFSS) Cognitive Decline Module at national and New York State levels. Leveraging large population-based BRFSS cohorts of 298,944 participants nationwide and 30,366 participants in New York [13], we compared 16 machine learning algorithms and four end-to-end modeling pipelines under chronological train/validation/locked-test splits. We further identified key factors contributing to SCD using machine learning-based feature importance analyses, post-hoc logistic models (national: unweighted; New York: design-weighted), and training-only knowledge-graph summaries. Altogether, this study contributes a reusable computational framework for population-level SCD risk ranking and identifies demographic, behavioral, and health-related factors associated with SCD.

## 2. Materials and Methods

### 2.1. Study design and data sources

We conducted a retrospective, cross-sectional prediction study using pooled Behavioral Risk Factor Surveillance System (BRFSS) annual survey files from 2015 through 2024 [13]. BRFSS is a United States state-based telephone survey of non-institutionalized adults conducted by the Centers for Disease Control and Prevention (CDC) [2, 14]. Survey data and codebooks are publicly available from the CDC BRFSS website (https://www.cdc.gov/brfss/index.html). Raw annual XPORT files were converted to comma-separated values and harmonized across survey years, following practices for multi-year BRFSS integration. Two analysis cohorts were constructed and analyzed independently. The national cohort pooled all states in which the Cognitive Decline module was fielded (analytic n = 298,944), with 38,238 participants in the training set, 112,561 in the validation set, and 148,145 in the locked test set. The New York cohort comprised respondents with state Federal Information Processing Standards code 36 (analytic n = 30,366), with 15,705 participants in the training set, 7,166 in the validation set, and 7,495 in the locked test set.

### 2.2. Study population and outcome definition

Eligible participants were adults aged ≥45 years who provided a valid response to the SCD screener in years and states with the Cognitive Decline module [15]. Standard BRFSS missing and refused codes were recoded to missing. The primary endpoint was SCD, coded 1 for “Yes” and 0 for “No” on the BRFSS confusion or memory loss item (CIMEMLOS for 2015-2022; CIMEMLO1 from 2023 onward) [13]. Respondents with “Don’t know,” “Refused,” or missing screener values were excluded. Unless otherwise noted, BRFSS categorical predictors retained native codebook coding. In particular, difficulty deciding (DECIDE) was retained as 1 = Yes and 2 = No and was not recoded to 0/1 for modeling or GLMs. Many other Yes/No health items use the same 1 = Yes, 2 = No codebook scale. In GLMs that retain this coding, an aOR < 1 comparing No versus Yes indicates higher SCD odds among respondents who answered Yes.

### 2.3. Predictor variables

We considered up to 58 harmonized BRFSS predictors covering demographics, health status, chronic conditions, functional difficulties, lifestyle, healthcare access, and BMI-related measures. Predictors retained native codebook coding, and year-specific availability followed BRFSS module fielding. This candidate set entered train-fitted preprocessing and nested LASSO selection; downstream supervised models and post-hoc association analyses used the resulting LASSO- stable subset. The number of selected predictors is reported in the Results.

### 2.4. Temporal partitioning

We applied a fixed chronological partition: training (2015-2019) for feature selection and model fitting; validation (2020-2022) for algorithm selection, calibration, and threshold tuning; and test (2023-2024) for a single locked performance evaluation. The test partition was not used for imputation, scaling, feature selection, algorithm choice, calibration, threshold selection, or interpretability analysis. For New York, the 2022 SCD module was not fielded; the New York validation set spans 2020-2021 only.

### 2.5. Data preprocessing and batch-effect adjustment

Preprocessing used train-fitted parameters throughout. (1) Cohort-level batch effect correction (before LASSO): survey year was treated as a batch variable. On the training partition only (2015- 2019), we median-imputed missing values, then applied ComBat-lite correction [16] by subtracting (μ_year - μ_global) for each feature. The training-fitted median imputer was applied to validation and test. Batch offsets were applied only for training survey years; validation and test rows received imputation only. (2) Model pipelines: Nested LASSO and classifiers were implemented as scikit-learn Pipelines [17] or tidymodels recipes [18] with median imputation and z-score standardization fit on training (or inner-fold training) data only. All supervised steps used the LASSO-stable predictor subset. Training-split PCA quality-control plots were generated before and after batch effect correction. Synthetic Minority Oversampling Technique (SMOTE) [19] was applied only inside cross-validation training folds when comparing the sixteen algorithms, scoring LASSO outer folds with a reference classifier, and evaluating ensemble candidates. Validation-based model selection, final model fitting, isotonic calibration, and locked test evaluation used the original class distribution (no SMOTE). Post-hoc interpretable GLMs used median imputation on LASSO features only, with no batch effect correction or SMOTE, so odds ratios retain standard epidemiologic interpretation.

### 2.6. Feature selection

We used L1-penalized logistic regression (LASSO) [20] with nested stratified group k-fold cross-validation by survey year on the batch effect-corrected training partition. The inverse regularization strength (C) (C = 1/lambda); smaller (C) implies stronger L1 shrinkage) was tuned by inner cross-validation. Within each outer fold, LASSO was refit at the selected (C), retaining at most 30 non-zero features. Predictors selected in at least 50% of outer folds formed the LASSO- stable set for downstream analyses; if fewer than five met this criterion, a fallback fit on the full training partition used the median optimal (C). Fold-wise median imputation and z-scoring were learned on fold-training data only. SMOTE was not used for feature selection. Python (scikit-learn) and R (glmnet) (C = 1/lambda) grids are documented in the repository.

### 2.7. Candidate classifiers and implementations

We implemented six analytic pipelines. Pipelines 1-4 were end-to-end SCD prediction workflows in Python and R (single-model or soft-voting). Within each prediction pipeline, the same sixteen supervised classifiers were evaluated on the LASSO-selected features; single-model pipelines retained the top validation performer, and soft-voting pipelines averaged predicted probabilities from the top three classifiers. Pipelines 5 and 6 were post-hoc training-only knowledge-graph analyses used for interpretation only. Classifier names and package versions are listed in the Software section. Sixteen algorithms were compared on LASSO-selected predictors using five-fold stratified group cross-validation by survey year (SMOTE restricted to training folds, as above). The single-model workflow retained the classifier with the best validation ROC-AUC among the sixteen candidates, and the soft-voting workflow averaged predicted class probabilities from the three top validation-ranked classifiers; both were refitted on all training data without SMOTE. All four Python/R prediction workflows used the same temporal splits, batch-effect correction, nested LASSO, validation isotonic calibration, and validation Youden J threshold [21]. Validation permutation importance and SHAP summaries were used for interpretability of the primary reporting pipelines.

### 2.8. Primary prediction pipeline designation and downstream reporting

To preserve a true locked test evaluation, primary reporting pipelines were prespecified before inspecting locked-test metrics. For the national cohort, the primary pipeline was the R tidymodels single-model workflow; for New York, the primary pipeline was the Python single-model workflow. These choices were made a priori to demonstrate cross-language replication rather than to maximize test performance. Within each pipeline, the final classifier (or soft-voting member set) was selected solely by validation ROC-AUC after refitting on the full training partition without SMOTE. Soft-voting Python and R pipelines were retained as sensitivity benchmarks. After validation-based selection, calibration, and threshold tuning, each of the four pipelines was scored once on the locked 2023-2024 test set. We do not designate a winner by highest test ROC-AUC.

### 2.9. Final model training, calibration, and test evaluation

Each prediction workflow used the same final steps after validation-based model choice. For single-model prediction pipelines, the validation-winning algorithm was refitted once on all training data without SMOTE. Predicted probabilities were calibrated with isotonic regression fit on validation only, then applied to test without refitting the calibrator. On those validation scores, we selected the probability threshold that maximized Youden’s J (sensitivity + specificity − 1) and froze that threshold for a single locked test evaluation. In soft-voting pipelines, the top three validation algorithms were refitted, their positive-class probabilities were averaged, and the same calibration and threshold rules were applied. On the locked 2023-2024 test partition we report discrimination and classification metrics at the frozen threshold: ROC-AUC, PR-AUC, accuracy, balanced accuracy ([sensitivity + specificity]/2), sensitivity, specificity, F1, and Brier score. Unless noted otherwise, 95% confidence intervals used nonparametric bootstrap resampling of test rows with replacement (B = 1,000; percentile intervals). Calibration was summarized with reliability curves (mean predicted probability versus observed SCD fraction within bins) and the Brier score. Machine learning models did not use BRFSS survey weights. Permutation importance and SHAP summaries [22] were computed on validation predictions only and were not used for model selection.

### 2.10. Statistical analysis and post-hoc inference

Descriptive SCD prevalences were unweighted unless otherwise stated. For pooled descriptive comparisons, we report binomial proportions with Wilson 95% confidence intervals and compare groups with chi-square tests (or two-sample tests of proportions); two-sided P < 0.05 was considered statistically significant for these descriptive contrasts. After locked test evaluation, we fit post-hoc logistic regression models of SCD on the LASSO-stable predictor set using training data only. National models used unweighted logistic regression because training-year state coverage was sparse. New York models used design-weighted survey logistic regression (svyglm) with BRFSS design variables (strata, primary sampling unit, and survey weight); rows with missing design variables were excluded from weighted fits. We report adjusted odds ratios (aORs) with 95% confidence intervals. Wald P-values for predictor coefficients were adjusted across features within each cohort model using the Benjamini-Hochberg false discovery rate (FDR), with q < 0.05 denoting statistical significance [23]. Predictors retained native BRFSS codebook coding. These GLMs were intended as associational complements to the prediction pipelines, not as replacements for SHAP or permutation rankings.

### 2.11. Knowledge-graph association analysis

For each cohort, we built a directed graph with an SCD node and predictors as nodes using training data only. In the data-only layer, edges combined four association sources: point-biserial (marginal) correlation with SCD, univariate logistic association, partial correlation of each predictor with SCD controlling for other predictors, and pairwise partial correlation among predictors (|r| threshold applied in software). When multiple sources produced the same directed edge, edge weights were summed. Each predictor received a composite association score: 0.25 × PageRank [24] + 0.15 × personalized PageRank toward SCD [25] + 0.15 × path proximity + 0.35 × normalized direct-edge strength to SCD + 0.10 × betweenness [26], with shortest-path distance defined as 1 / (weight + ε) [27, 28]. This analysis summarizes statistical co-structure on training data only and is not causal inference. A complementary ML-integrated layer, run after post-hoc GLMs, built a similar graph on LASSO-selected predictors plus SCD. Edges integrated training- split GLM odds ratios (national unweighted; New York design-weighted), validation permutation importance from the Python single-model pipeline, pairwise partial correlations on training data, and predefined health-topic priors. Duplicate edges were summed and node scores used the same composite network metric as the data-only layer. This layer combines survey regression, ML importance, partial correlations, and health-topic priors for hypothesis-generating interpretation only; proximity in the graph should not be read as a biological mechanism.

### 2.12. Software implementation and reproducibility

Analyses used Python 3.12 and R 4.5.3 with fixed random seeds (42). BRFSS data are publicly available from the CDC. Analysis code, processed cohort files, and end-to-end pipeline scripts are available at https://github.com/truong128/brfss-scd-ml-benchmark. The repository implements harmonized cohort construction; train-fitted preprocessing and nested LASSO; classifier benchmarking with validation-based selection, isotonic calibration, and Youden thresholding; locked test scoring with bootstrap intervals; and post-hoc GLM/knowledge-graph exports. Publication figures can be regenerated from scored pipeline outputs. The six pipelines were: (1) Python single-model ML; (2) Python voting-soft ML; (3) R tidymodels voting-soft ML; (4) R tidymodels single-model ML; (5) ML-integrated knowledge-graph association analysis; and (6) data-only knowledge-graph association analysis. Pipelines 1-4 compared the same sixteen classifiers spanning linear, kernel, tree ensemble, nearest-neighbor, discriminant, naive Bayes, and neural-network families. Pipelines 5 and 6 did not use classifier benchmarking. Python and R package versions are pinned in the repository setup scripts.

## 3. Results

### 3.1. Rising SCD prevalence across national and New York survey years

We included adults aged 45 years or older with a valid SCD response from BRFSS Cognitive Decline module years 2015-2024 (national n = 298,944; New York n = 30,366; Table 1; Figure 1A). Chronological split sizes follow the design in Methods. Table 1 and Supplementary Figure S1 show unweighted SCD prevalence and sample size by year. Nationally, training-year SCD prevalence was 11.2% with only one to seven states contributing each year (year-specific range 9.3%-12.0%). Validation covered 2020-2022, and locked test years 2023-2024 had broader state coverage, with SCD prevalence 16.9% in 2023 and 17.2% in 2024 (pooled 17.0%). In New York, training prevalence was 10.3% (range 9.2%-11.6%), validation covered 2020-2021 only because the module was not fielded in 2022, and test prevalence was 16.4% in 2023 and 15.5% in 2024 (pooled 15.8%).

**Figure 1.**
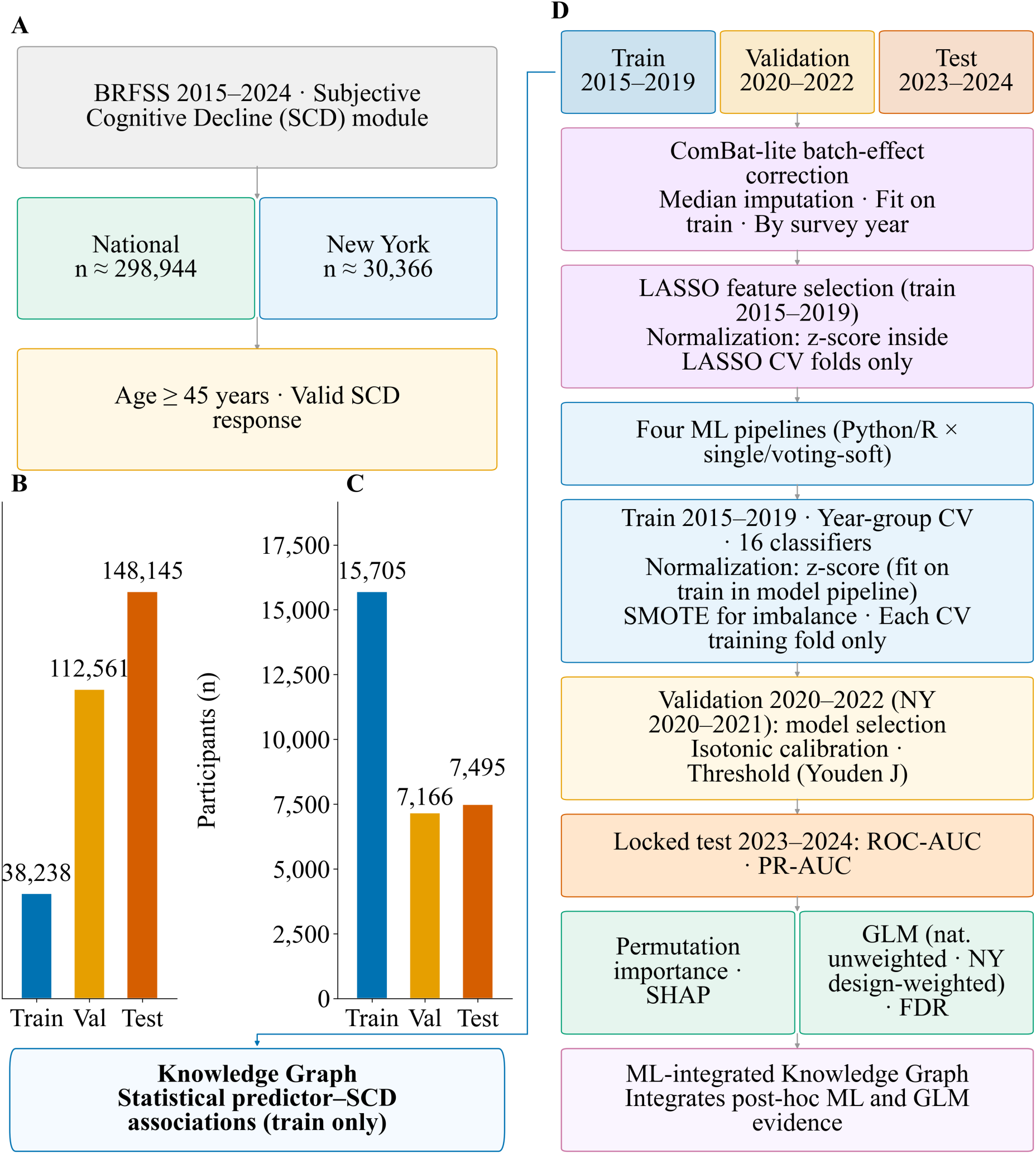
Study design and analytic workflow. (A) BRFSS 2015-2024 data sources, national and New York analytic cohorts, and inclusion criteria (age ≥45 years; valid SCD response). (B, C) Participants by temporal split for the national (B) and New York (C) cohorts (train, validation, test; New York validation is 2020-2021 because the module was not fielded in 2022). (D) Locked temporal splits and end-to-end workflow: train-fit median imputation and ComBat-lite batch correction by survey year; nested LASSO feature selection with z-score normalization inside LASSO CV folds; four prediction pipelines (1-4; Python/R × single/voting-soft) with z-score normalization in the model processing pipeline and SMOTE in train CV folds only; validation algorithm selection, isotonic calibration, and classification threshold (Youden J); locked test evaluation (2023-2024); post-hoc permutation importance, SHAP, and GLMs (national unweighted; New York design-weighted; FDR, BH); ML-integrated knowledge graph (pipeline 5). Data-only knowledge graph (pipeline 6): statistical predictor-SCD associations on the training split only, branching from the Train partition (box below panels B-C).

**Table 1.**
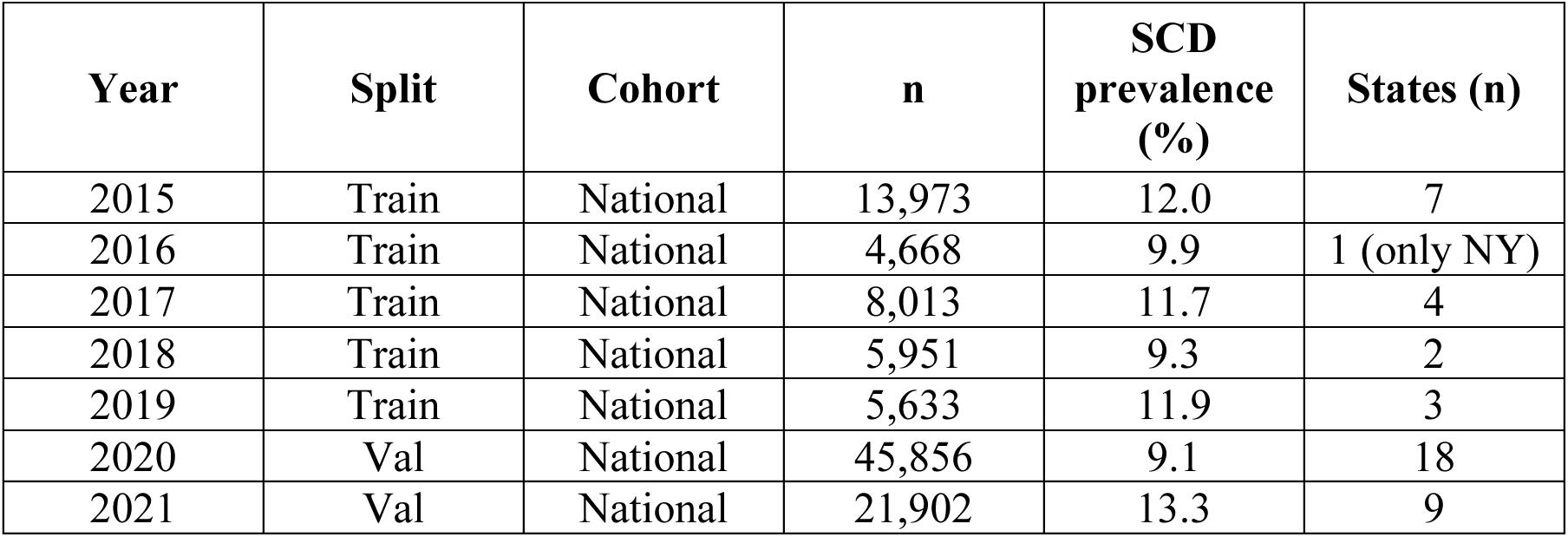
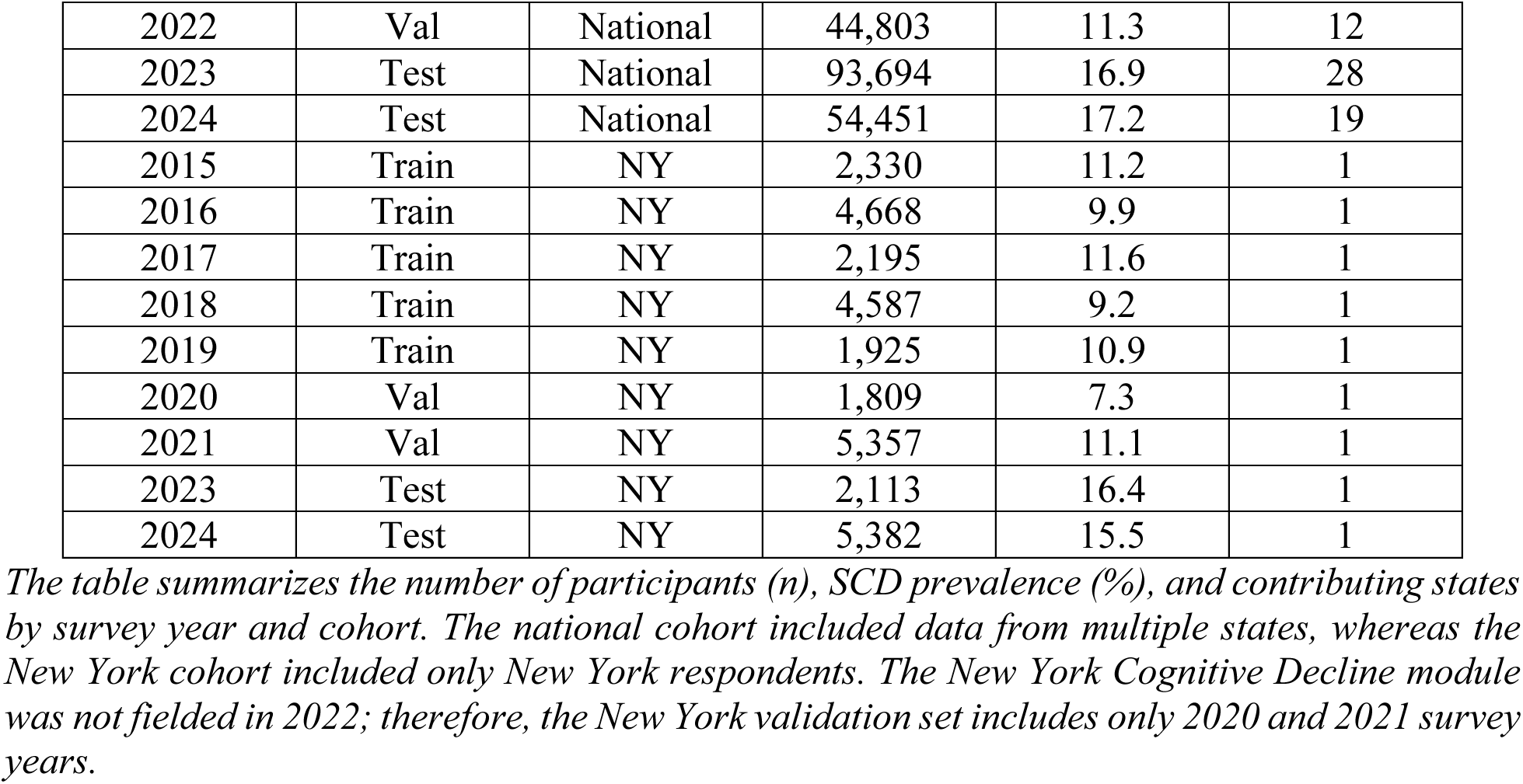
Analytic sample size and unweighted SCD prevalence by survey year.

| Year | Split | Cohort | n | SCD prevalence (%) | States (n) |
| --- | --- | --- | --- | --- | --- |
| 2015 | Train | National | 13,973 | 12.0 | 7 |
| 2016 | Train | National | 4,668 | 9.9 | 1 (only NY) |
| 2017 | Train | National | 8,013 | 11.7 | 4 |
| 2018 | Train | National | 5,951 | 9.3 | 2 |
| 2019 | Train | National | 5,633 | 11.9 | 3 |
| 2020 | Val | National | 45,856 | 9.1 | 18 |
| 2021 | Val | National | 21,902 | 13.3 | 9 |
| 2022 | Val | National | 44,803 | 11.3 | 12 |
| 2023 | Test | National | 93,694 | 16.9 | 28 |
| 2024 | Test | National | 54,451 | 17.2 | 19 |
| 2015 | Train | NY | 2,330 | 11.2 | 1 |
| 2016 | Train | NY | 4,668 | 9.9 | 1 |
| 2017 | Train | NY | 2,195 | 11.6 | 1 |
| 2018 | Train | NY | 4,587 | 9.2 | 1 |
| 2019 | Train | NY | 1,925 | 10.9 | 1 |
| 2020 | Val | NY | 1,809 | 7.3 | 1 |
| 2021 | Val | NY | 5,357 | 11.1 | 1 |
| 2023 | Test | NY | 2,113 | 16.4 | 1 |
| 2024 | Test | NY | 5,382 | 15.5 | 1 |

*The table summarizes the number of participants (n), SCD prevalence (%), and contributing states by survey year and cohort. The national cohort included data from multiple states, whereas the New York cohort included only New York respondents. The New York Cognitive Decline module was not fielded in 2022; therefore, the New York validation set includes only 2020 and 2021 survey years.*

Test-year prevalence was higher than training-year prevalence in both cohorts (Supplementary Figure S1), supporting the locked 2023-2024 holdout described in Methods. Pooled SCD prevalence was 13.6% nationally versus 11.4% in New York among adults aged 45 years or older (Supplementary Figure S2; P < 0.001). Prevalence also varied by race and ethnicity (Supplementary Figure S3) and showed only small sex differences (Supplementary Figure S4).

### 3.2. Stable LASSO features and validation-guided algorithm selection

Nested LASSO on training data selected 24 stable predictors in the national cohort (outer cross-validated ROC-AUC 0.796) and 30 in New York (outer ROC-AUC 0.789; Tables 2A-B; Figure 2). Sixteen algorithms were then compared within each end-to-end prediction workflow using the LASSO-stable predictor subset (Figure 3). Nationally, gradient boosting ranked highest on validation (0.794; Table 3A; Figure 3A). In New York, AdaBoost had the highest validation ROC-AUC (0.815; Table 3B; Figure 3B), although linear models ranked higher during training cross-validation alone (Figure 3C-D). Soft-voting ensemble benchmarks on training-fold cross-validation were essentially tied with the best single algorithm and did not meaningfully outperform it (Figure 3E-F).

**Table 2A.** LASSO feature stability (National).

| Rank | Feature | Selection frequency | Included in final model |
| --- | --- | --- | --- |
| 1 | age group | 1.00 | Yes |
| 2 | arthritis | 1.00 | Yes |
| 3 | blindness | 1.00 | Yes |
| 4 | COPD | 1.00 | Yes |
| 5 | depression disorder | 1.00 | Yes |
| 6 | difficulty being alone | 1.00 | Yes |
| 7 | difficulty deciding | 1.00 | Yes |
| 8 | difficulty walking | 1.00 | Yes |
| 9 | flu shot | 1.00 | Yes |
| 10 | general health | 1.00 | Yes |
| 11 | mental health days | 1.00 | Yes |
| 12 | sex | 1.00 | Yes |
| 13 | arthritis disability | 0.80 | Yes |
| 14 | average drinks | 0.80 | Yes |
| 15 | BMI category | 0.80 | Yes |

*Top 15 of 24 stable features are shown (selected in $\geq 50\%$ of nested outer folds) in the national training cohort. Nested outer cross-validated ROC-AUC = 0.796 (SD 0.010); median penalty C = 100 (R tidymodels implementation). All listed features were included in the final model.*

**Table 2B.** LASSO feature stability (New York).

| Rank | Feature | Selection frequency | Included in final model |
| --- | --- | --- | --- |
| 1 | age group | 1.00 | Yes |
| 2 | arthritis | 1.00 | Yes |
| 3 | average drinks | 1.00 | Yes |
| 4 | blindness | 1.00 | Yes |
| 5 | blood pressure medication | 1.00 | Yes |
| 6 | depression disorder | 1.00 | Yes |
| 7 | difficulty being alone | 1.00 | Yes |
| 8 | difficulty deciding | 1.00 | Yes |
| 9 | difficulty walking | 1.00 | Yes |
| 10 | flu shot | 1.00 | Yes |
| 11 | general health | 1.00 | Yes |
| 12 | Hispanic | 1.00 | Yes |
| 13 | mental health days | 1.00 | Yes |
| 14 | physical health days | 1.00 | Yes |
| 15 | routine checkup | 1.00 | Yes |

*Top 15 of 30 stable features are shown (selected in $\geq 50\%$ of nested outer folds) in the New York training cohort. Nested outer cross-validated ROC-AUC = 0.789 (SD 0.025); median penalty C = 0.01 (Python implementation). All listed features were included in the final model.*

**Figure 2.**
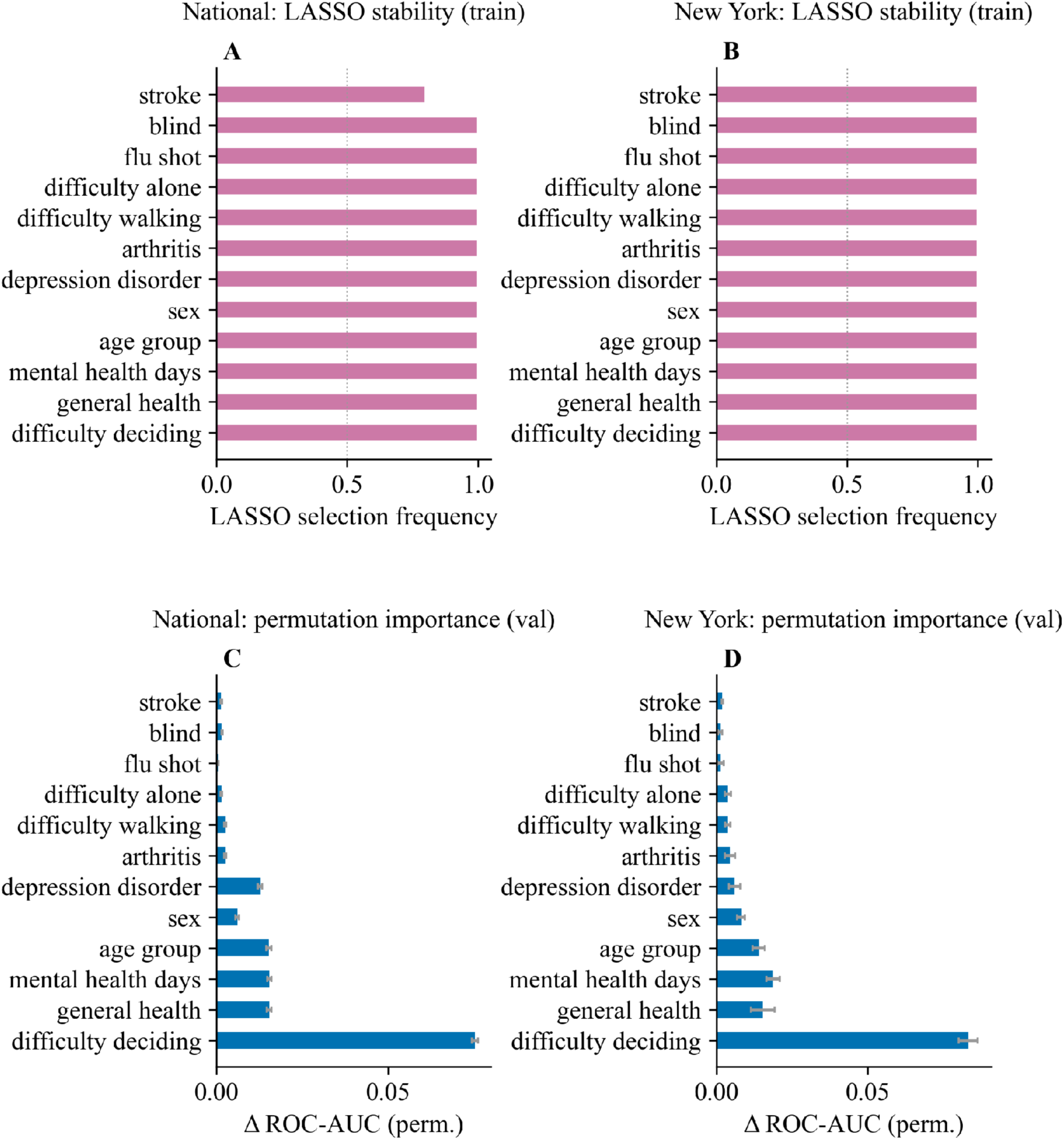
Feature selection and permutation importance. National left; New York right. (A, B) LASSO feature-selection stability on training data (top 12 features, aligned). (C, D) Validation-set permutation importance for the lead single-model pipelines (national: R gradient boosting; New York: Python AdaBoost).

**Figure 3.**
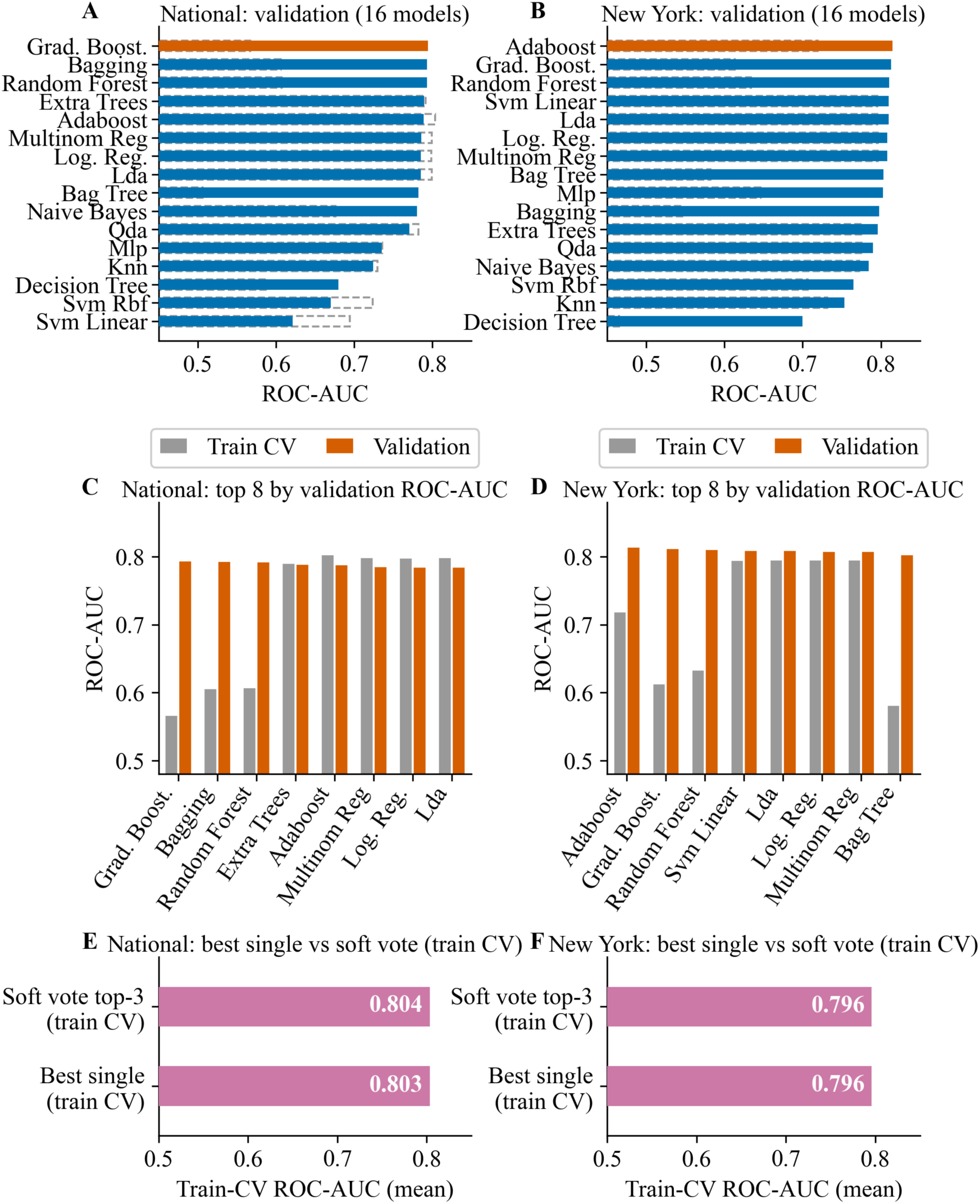
Classifier and ensemble comparison. National cohort results are shown in the left panels (A, C, E; R tidymodels single-model pipeline), and New York cohort results are shown in the right panels (B, D, F; Python single-model pipeline). (A, B) Validation ROC-AUC for sixteen candidate classifiers (solid bars); gray dashed outlines show the corresponding training-fold cross-validation mean ROC-AUC. The orange bar marks the validation-selected best single algorithm in each cohort. (C, D) Training-fold cross-validation versus validation ROC-AUC for the top eight classifiers by validation ROC-AUC (legend: Train CV, Validation). (E, F) Best single algorithm versus soft-vote top-3 ensemble under matched training-fold cross-validation mean ROC-AUC.

**Table 3A.**
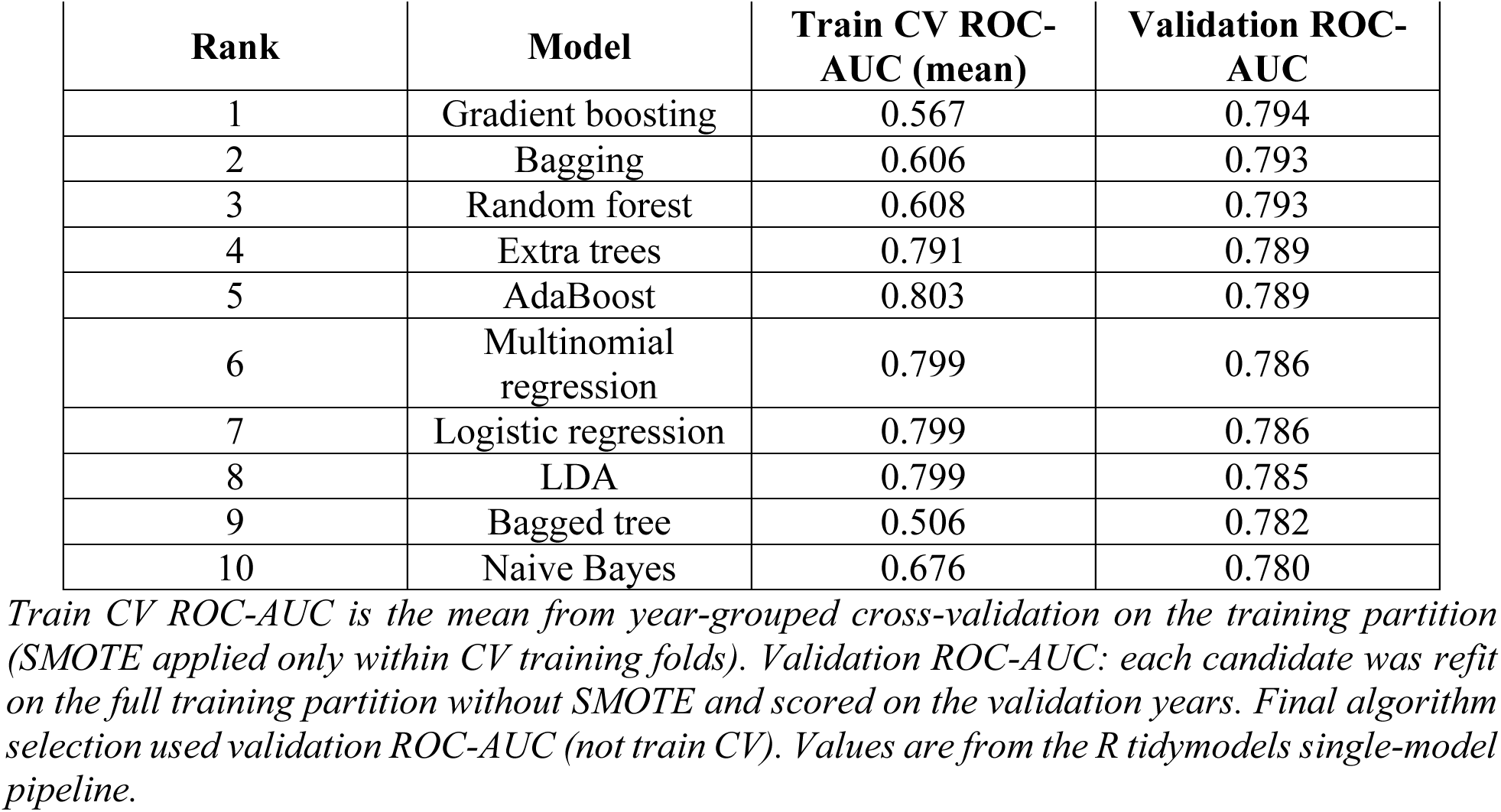
Top ten classifiers by validation ROC-AUC (National).

**Table 3B.**
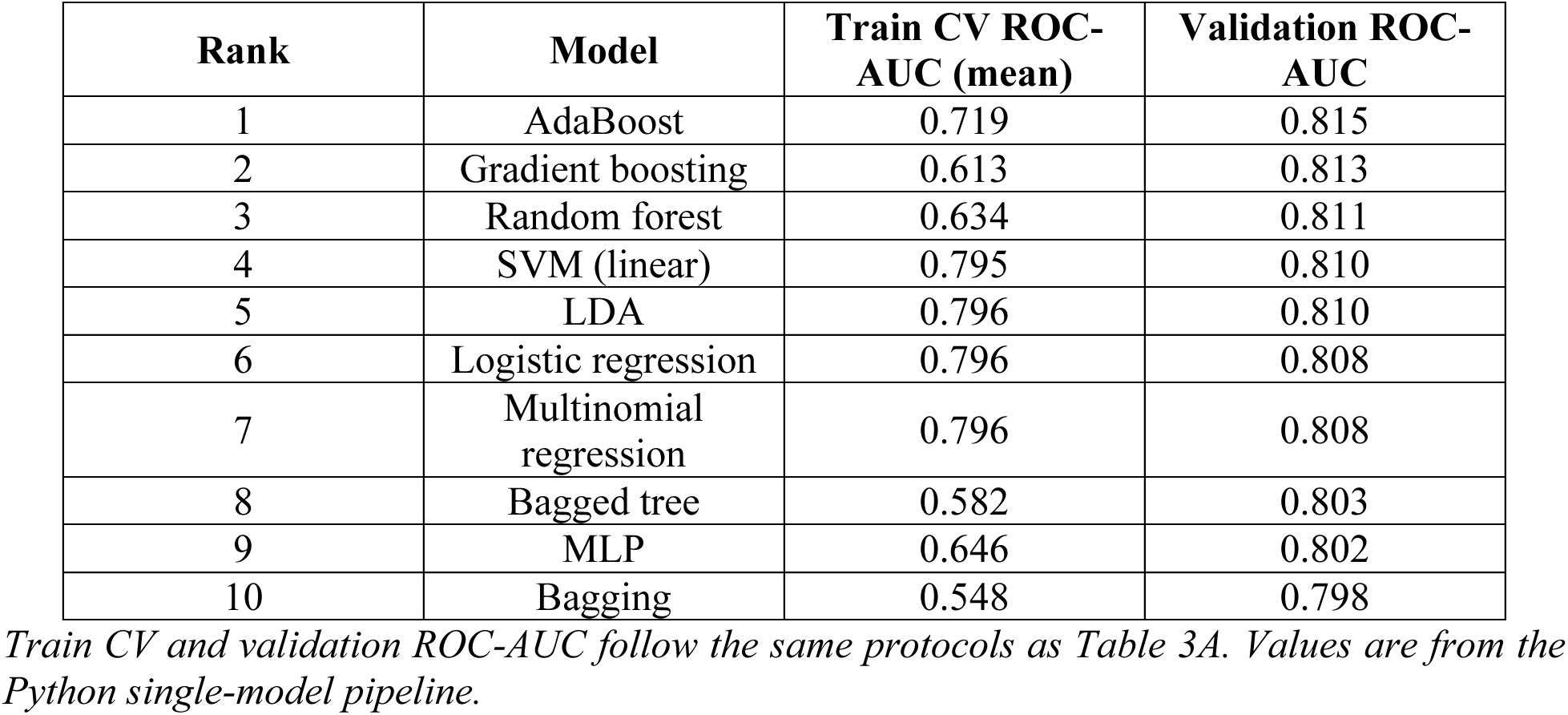
Top ten classifiers by validation ROC-AUC (New York).

| Rank | Model | Train CV ROC-AUC (mean) | Validation ROC-AUC |
| --- | --- | --- | --- |
| 1 | AdaBoost | 0.719 | 0.815 |
| 2 | Gradient boosting | 0.613 | 0.813 |
| 3 | Random forest | 0.634 | 0.811 |
| 4 | SVM (linear) | 0.795 | 0.810 |
| 5 | LDA | 0.796 | 0.810 |
| 6 | Logistic regression | 0.796 | 0.808 |
| 7 | Multinomial regression | 0.796 | 0.808 |
| 8 | Bagged tree | 0.582 | 0.803 |
| 9 | MLP | 0.646 | 0.802 |
| 10 | Bagging | 0.548 | 0.798 |

*Train CV and validation ROC-AUC follow the same protocols as Table 3A. Values are from the Python single-model pipeline.*

### 3.3. Concordant locked-test ranking across four Python/R pipelines

We compared the four prediction pipelines on the locked 2023-2024 test set (Table 4; Figure 4). All four pipelines performed similarly and showed ability to rank who was more likely to report SCD. By ROC-AUC (overall ranking quality), national scores were 0.769-0.770 and New York scores were 0.754-0.762; New York intervals were wider because fewer test respondents were available. We also report PR-AUC, which focuses more on correctly identifying the SCD-positive group (Figure 4B): 0.483-0.494 nationally and 0.439-0.456 in New York. These PR-AUC numbers are below 0.5, but that does not mean the models were worse than guessing. SCD was reported by only about 16-17% of test respondents (Table 5), so a model that ignored all health predictors and just reflected that low rate would score near 0.16-0.17 on PR-AUC. Against that simple reference, the observed PR-AUC values are substantially higher. Soft-voting pipelines did not clearly beat the best single model. Detailed reporting therefore uses the primary single-model pipelines (R gradient boosting nationally; Python AdaBoost in New York), with soft-voting kept as a sensitivity check.

**Figure 4.**
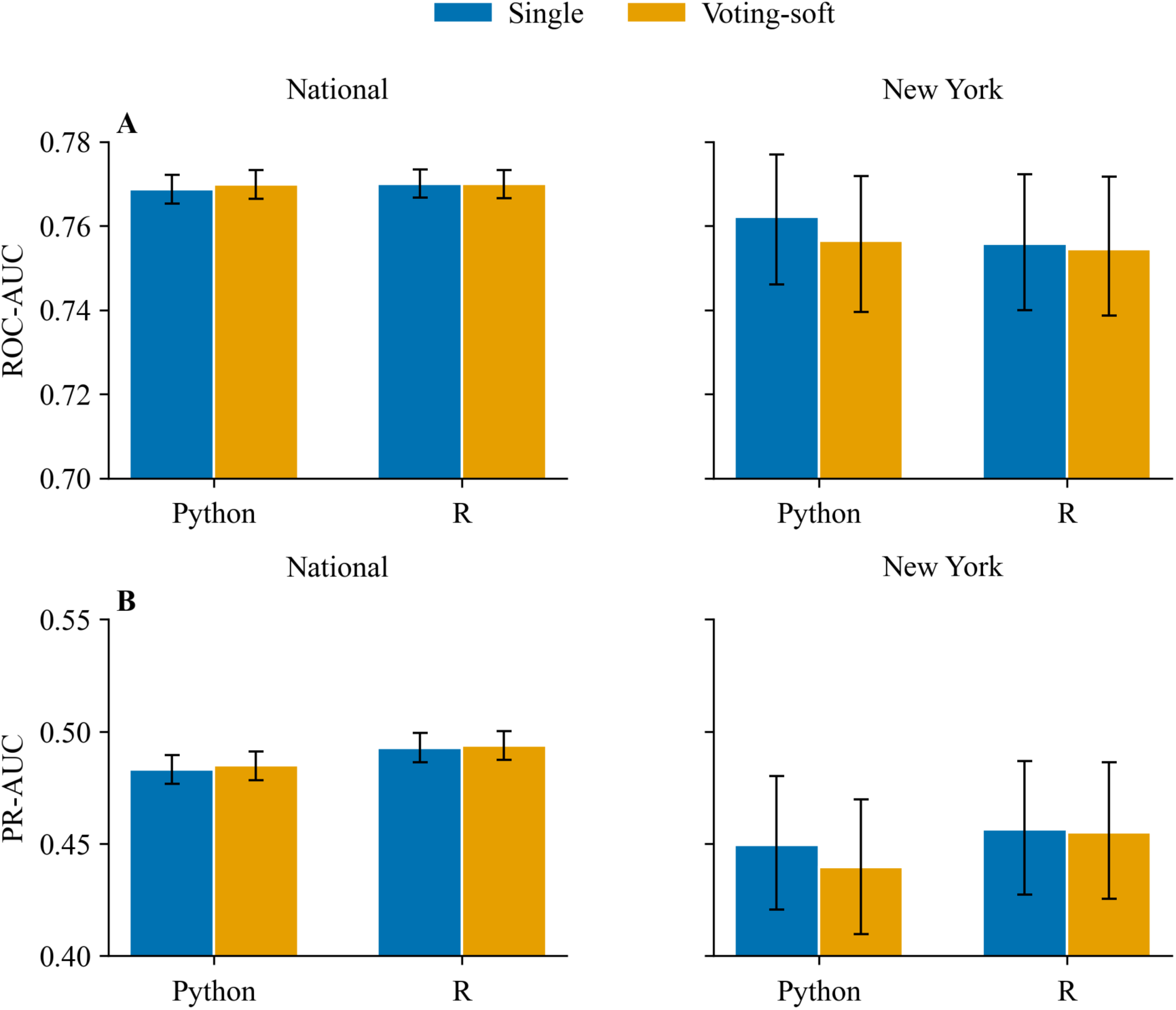
Locked test-set performance across four prediction pipelines (2023-2024). (A) Test ROC-AUC for the national (left) and New York (right) cohorts. (B) Test PR-AUC for the national (left) and New York (right) cohorts. Within each panel, grouped bars compare Python and R; blue = single-model pipeline and orange = soft-voting pipeline. Error bars are nonparametric bootstrap 95% confidence intervals (B = 1,000). For PR-AUC (B), a model that ignored predictors would score near the test-set SCD prevalence (∼16-17%; Table 5), not near 0.5.

**Table 4.** Locked test-set ROC-AUC by prediction pipeline (2023-2024).

| Prediction pipeline | National model | National ROC-AUC (95% CI) | NY model | NY ROC-AUC (95% CI) |
| --- | --- | --- | --- | --- |
| 1. Python single | Gradient boosting | 0.769 (0.765-0.772) | AdaBoost | 0.762 (0.746-0.777) |
| 2. Python soft-voting | Soft vote | 0.770 (0.766-0.773) | Soft vote | 0.757 (0.740-0.772) |
| 3. R tidymodels single | Gradient boosting | 0.770 (0.767-0.773) | AdaBoost | 0.756 (0.740-0.772) |
| 4. R tidymodels soft-voting | Soft vote | 0.770 (0.767-0.773) | Soft vote | 0.754 (0.739-0.772) |

*Values are locked-test ROC-AUC with nonparametric bootstrap 95% confidence intervals ( $B = 1,000$ ). Primary reporting pipelines: R tidymodels single (national) and Python single (New York); soft-voting rows are sensitivity benchmarks. Soft vote = probability average of the top-3 validation-ranked classifiers.*

**Table 5.** Locked test-set performance of the primary reporting pipelines (2023-2024).

| **Metric** | **National (R gradient boosting)** | **New York (Python AdaBoost)** |
| --- | --- | --- |
| Test n | 148,145 | 7,495 |
| ROC-AUC (95% CI) | 0.770 (0.767-0.773) | 0.762 (0.746-0.777) |
| PR-AUC (95% CI) | 0.493 (0.486-0.499) | 0.449 (0.420-0.480) |
| Accuracy (95% CI) | 0.779 (0.777-0.781) | 0.814 (0.804-0.822) |
| Balanced accuracy | 0.699 | 0.688 |
| Sensitivity (95% CI) | 0.578 (0.573-0.584) | 0.505 (0.476-0.533) |
| Specificity (95% CI) | 0.820 (0.818-0.822) | 0.871 (0.863-0.880) |
| F1 (95% CI) | 0.470 (0.466-0.475) | 0.461 (0.435-0.483) |
| Brier score (95% CI) | 0.118 (0.116-0.119) | 0.112 (0.106-0.119) |
| Threshold (validation-tuned) | 0.126 | 0.129 |
| Test prevalence | 17.0% | 15.8% |

### 3.4. Primary pipelines: discrimination, calibration, and year-stable scores

Table 5 summarizes locked-test performance for the primary reporting pipelines after validation-based algorithm choice, calibration, and threshold tuning. Nationally (R gradient boosting; n = 148,145; test prevalence 17.0%), test ROC-AUC was 0.770 (95% CI 0.767-0.773), accuracy 0.779, sensitivity 0.578, specificity 0.820, and Brier score 0.118 at threshold 0.126 (Figure 5A,C). In New York (Python AdaBoost; n = 7,495; test prevalence 15.8%), test ROC-AUC was 0.762 (95% CI 0.746-0.777), accuracy 0.814, sensitivity 0.505, specificity 0.871, and Brier score 0.112 at threshold 0.129 (Figure 5B,D). At these Youden-optimized cutoffs, many SCD-positive respondents were still missed (about half in New York; about 42% nationally), while specificity remained relatively high. The Python soft-voting sensitivity pipeline was similar (0.770 nationally; 0.757 in New York; Supplementary Figure S5).

**Figure 5.**
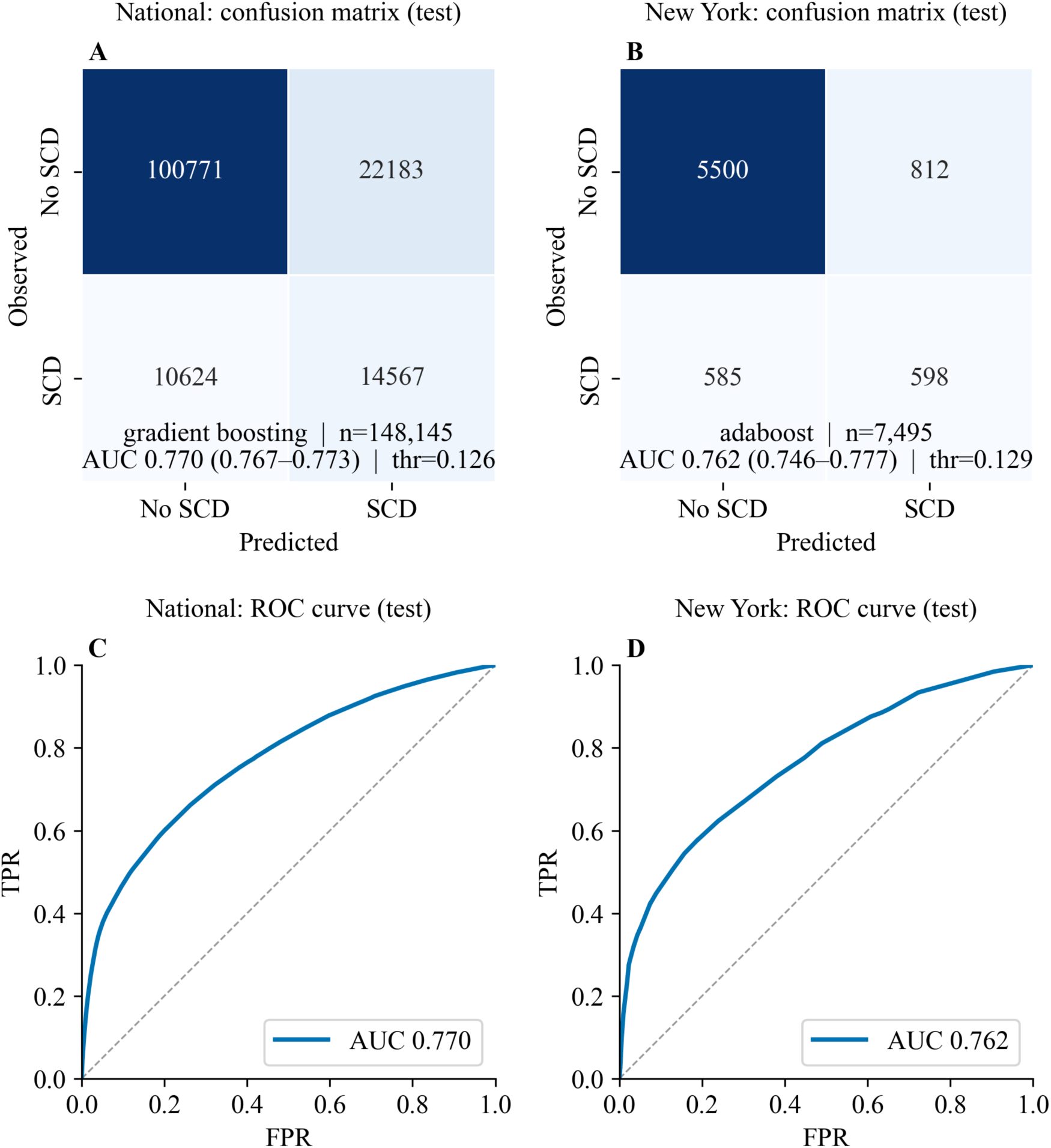
Locked test-set discrimination of the primary reporting pipelines (2023-2024). National results are shown on the left (A, C; R tidymodels gradient boosting) and New York results on the right (B, D; Python AdaBoost). (A, B) Confusion matrices at the validation-tuned probability threshold, with in-panel footnotes for model, sample size, test ROC-AUC (95% CI), and threshold. (C, D) ROC curves from the same locked test predictions. Additional metrics, including PR-AUC, are reported in Table 5.

On the locked test set, predicted probabilities agreed reasonably with observed SCD rates (Figure 6A-B; Brier 0.118 nationally and 0.112 in New York; Table 5), and scores were higher for SCD- positive than SCD-negative respondents (Figure 6C-D). The same separation held across survey years in validation and test data (Figure 7), even as unweighted SCD prevalence rose later. Thresholds and calibration should be revisited as new survey cycles accrue.

**Figure 6.**
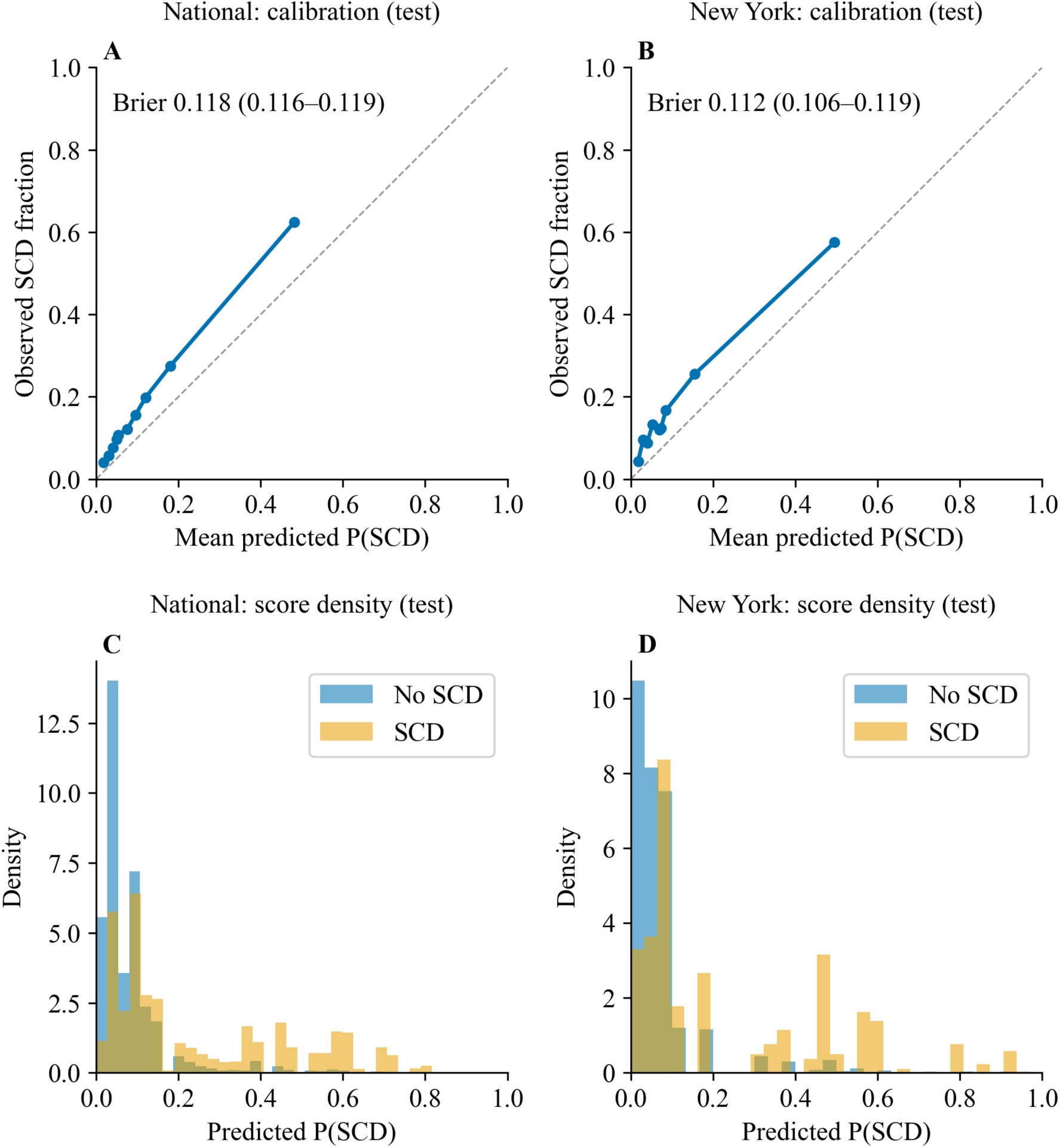
Calibration and score distributions on the locked test set (2023-2024). National left (A, C); New York right (B, D). (A, B) Calibration curves with in-panel Brier score (bootstrap 95% CI; Table 5). (C, D) Predicted P(SCD) densities by observed SCD status.

**Figure 7.**
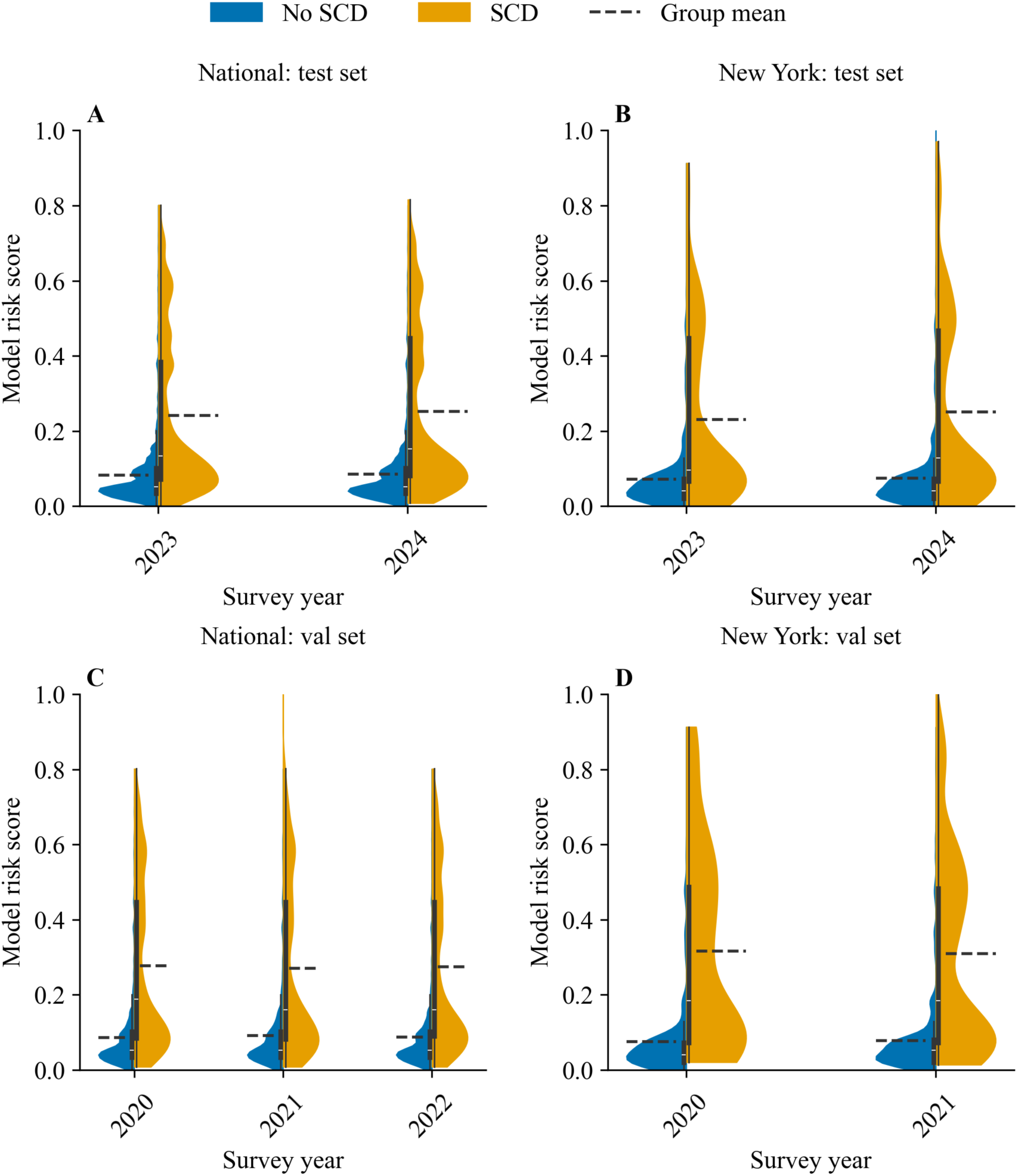
Model risk scores by survey year versus BRFSS-reported SCD. National left (A, C); New York right (B, D). (A, B) Test years 2023-2024. (C, D) Validation years (national 2020-2022; New York 2020-2021). Dashed lines are group means. SCD status is the evaluation label, not a model input.

### 3.5. Permutation, SHAP, and survey models converge on top predictors

We assessed predictor stability and importance on the primary reporting pipelines (Figure 2). Difficulty deciding showed the largest drop in validation ROC-AUC when shuffled (0.075 nationally and 0.083 in New York). Other contributors included general health, mental health days, age group, sex, and depression, but with much smaller effects.

SHAP summaries on validation data gave the same ranking: difficulty deciding contributed most to individual predictions in both cohorts (Figure 8A-B). Survey regression models on training data also ranked difficulty deciding highest. Respondents who reported no difficulty deciding (DECIDE = 2) had substantially lower odds of SCD than those who reported difficulty (DECIDE = 1; aOR approximately 0.13; q < 0.001; Table 6; Figure 8C-D). National models were unweighted; New York models used survey weights. Depression, general health, and functional limitation items were also associated with SCD after adjustment. These three approaches agreed on the same top predictors. Importance scores and odds ratios describe association and ranking, not proven cause.

**Figure 8.**
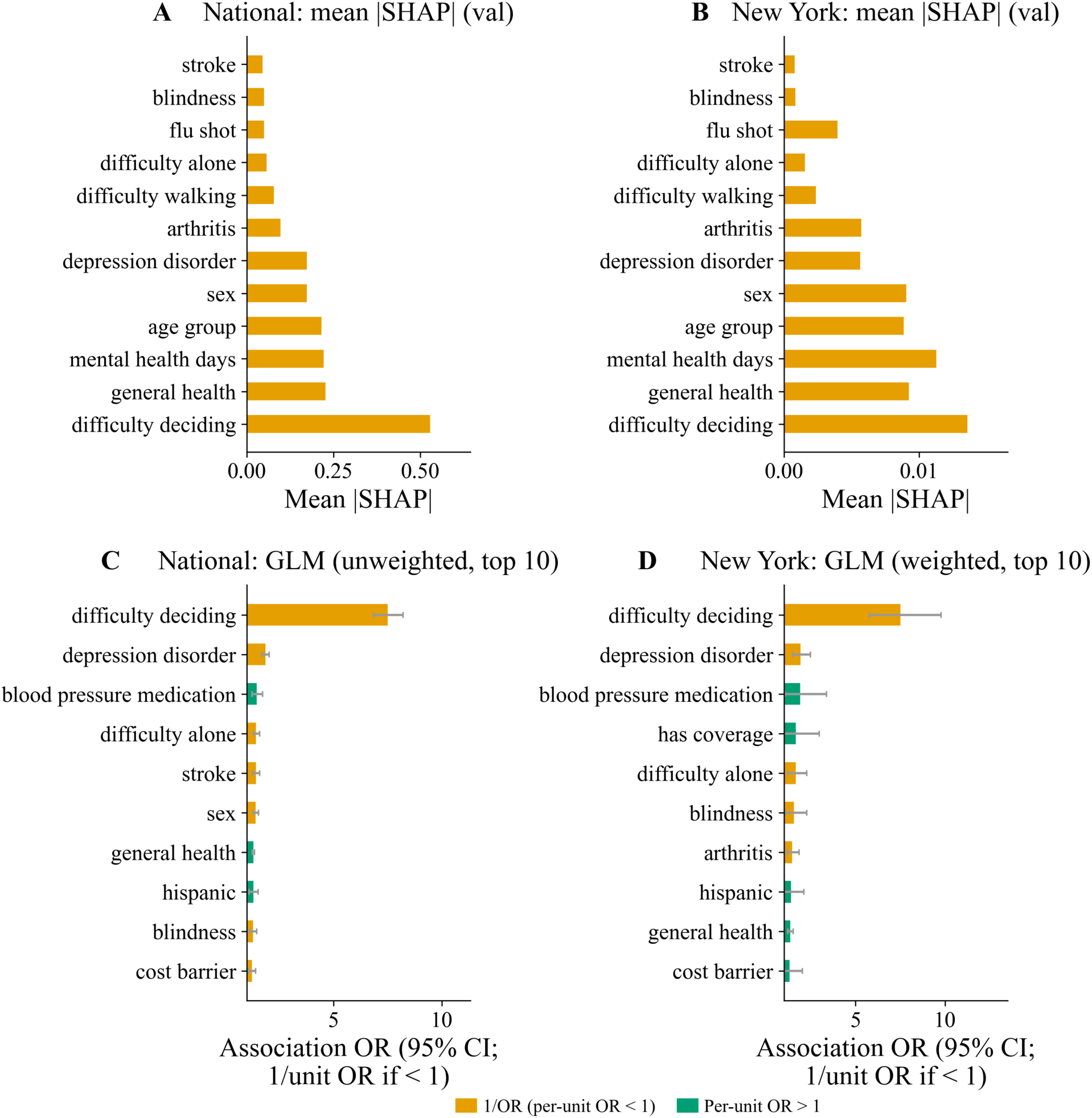
Model interpretability using SHAP and GLM associations. National left (A, C); New York right (B, D). (A, B) Mean absolute SHAP values on validation data for the primary reporting pipelines (national: R gradient boosting; New York: Python AdaBoost). (C, D) Top ten GLM associations on training data ranked by |log(OR)| (national: unweighted; New York: design-weighted). Bars are centered at OR = 1. For display only, modeled OR < 1 is plotted as 1/OR (orange) and OR > 1 as the OR itself (green); Table 6 reports untransformed aORs. For difficulty deciding (DECIDE = 1 “Yes”, 2 “No”), aOR ≈ 0.13 means “No” has lower SCD odds than “Yes”.

**Table 6.** GLM predictors with FDR q < 0.05 (LASSO feature set, training data).

**Panel A: National (unweighted GLM)**
| Predictor | aOR | 95% CI | q (BH) |
| --- | --- | --- | --- |
| difficulty deciding | 0.13 | 0.12-0.15 | <0.001 |
| depression disorder | 0.54 | 0.49-0.59 | <0.001 |
| general health | 1.30 | 1.25-1.36 | <0.001 |
| age group | 1.18 | 1.14-1.22 | <0.001 |
| sex | 0.71 | 0.65-0.78 | <0.001 |
| difficulty alone | 0.70 | 0.63-0.78 | <0.001 |
| stroke | 0.71 | 0.63-0.80 | <0.001 |
| mental health days | 1.02 | 1.01-1.02 | <0.001 |
| arthritis | 0.82 | 0.75-0.88 | <0.001 |
| flu shot | 0.83 | 0.76-0.89 | <0.001 |
| high blood pressure | 1.10 | 1.06-1.15 | <0.001 |
| blood pressure medication | 1.46 | 1.23-1.72 | <0.001 |
| physical health days | 0.99 | 0.98-0.99 | <0.001 |
| skin cancer | 0.81 | 0.74-0.89 | <0.001 |
| difficulty walking | 0.82 | 0.75-0.90 | <0.001 |
| blindness | 0.78 | 0.69-0.88 | <0.001 |
| bmi category | 0.91 | 0.87-0.95 | <0.001 |
| hispanic | 1.30 | 1.11-1.53 | 0.002 |
| cost barrier | 0.81 | 0.71-0.93 | 0.003 |
| arthritis disability | 0.84 | 0.75-0.94 | 0.004 |
| copd | 0.87 | 0.79-0.97 | 0.013 |
| average drinks | 1.03 | 1.01-1.06 | 0.018 |
| smoked 100 | 0.92 | 0.86-1.00 | 0.050 |

**Panel B: New York (svyglm, design-weighted)**
| Predictor | aOR | 95% CI | q (BH) |
| --- | --- | --- | --- |
| difficulty deciding | 0.13 | 0.10-0.17 | <0.001 |
| general health | 1.36 | 1.21-1.53 | <0.001 |
| depression disorder | 0.52 | 0.40-0.67 | <0.001 |
| arthritis | 0.68 | 0.54-0.86 | 0.008 |
| difficulty alone | 0.60 | 0.44-0.82 | 0.008 |
| age group | 1.13 | 1.03-1.23 | 0.037 |
| average drinks | 1.05 | 1.01-1.10 | 0.037 |
| diabetes | 0.84 | 0.75-0.96 | 0.037 |
| mental health days | 1.02 | 1.00-1.03 | 0.040 |
| blindness | 0.64 | 0.44-0.93 | 0.048 |
| smoked 100 | 0.77 | 0.62-0.96 | 0.048 |

*Unweighted logistic regression for the national cohort (panel A) and design-weighted survey logistic regression for New York (panel B) on training data. Predictors retain native BRFSS coding. For difficulty deciding, respondents answering No (DECIDE = 2) had substantially lower odds of SCD than those answering Yes (DECIDE = 1; aOR approximately 0.13). The same Yes = 1 / No = 2 scale applies to other Yes/No health items in the table; for those items, an aOR < 1 likewise indicates higher SCD odds for Yes than for No. National models were unweighted because training-year state coverage was sparse. P-values were adjusted using the Benjamini-Hochberg FDR procedure.*

### 3.6. Knowledge graphs place difficulty deciding nearest to SCD

We mapped statistical links among predictors and SCD using training data only (Figure 9). National and New York maps were similar. Difficulty deciding had the highest composite association score and the shortest direct link to SCD in both cohorts. Other close predictors included general health, depression, and functional limitation items. Longer or weaker links connected SCD to items such as physical activity, cost barriers, and employment. These maps describe associational co-structure rather than causal mechanisms. A second map added survey regression results, model importance, partial correlations, and health-topic groupings (Supplementary Figure S6). The same core predictors remained near SCD, with difficulty deciding still central in both cohorts, supporting concordance between the data-only and ML-integrated associational displays.

**Figure 9.**
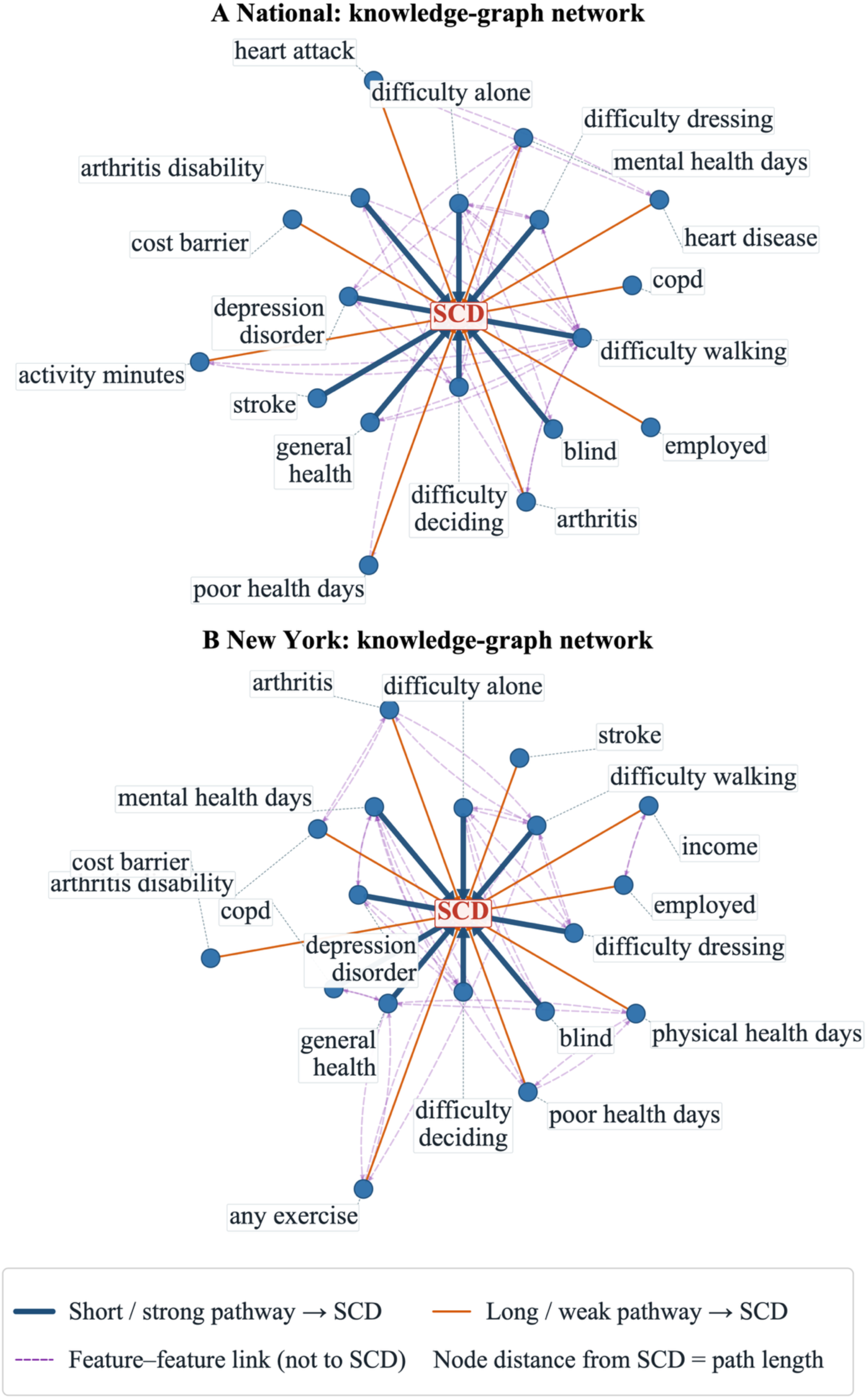
Data-only knowledge-graph associations with subjective cognitive decline (SCD). National (A) and New York (B). Networks were built from training-split harmonized covariates only (marginal and partial correlations plus univariate logistic associations; associational proximity only, not causation). The central red node is SCD; peripheral nodes are the 18 predictors with the highest composite association scores. Radial node distance reflects shortest-path graph distance to SCD. Dark blue solid lines, short/strong associational links to SCD (closest half by path distance); orange solid lines, longer/weaker links to SCD (farthest half); purple dashed lines, feature-feature links not terminating at SCD.

## 4. Discussion

We built and tested a reusable multi-language machine learning benchmark for self-reported SCD in large BRFSS cohorts from the United States and New York. Under chronological training (2015-2019), validation (national: 2020-2022; New York: 2020-2021), and a single locked 2023- 2024 test evaluation, primary pipelines reached test ROC-AUC about 0.76-0.77. Python and R implementations, and single-model versus soft-voting ensembles, performed similarly. Across feature selection, model explanations, survey regression, and network maps, the same core signals recurred: difficulty deciding, self-rated general health, and depression.

Most prior BRFSS work describes SCD prevalence and correlates rather than predicting SCD on future survey years. To our knowledge, previous studies [10–12] have not combined fixed year-based temporal validation at this scale with an exportable multi-language end-to-end pipeline. The national analytic sample of nearly 300,000 adults is much larger than many clinical or imaging SCD machine-learning studies and relies on questions already collected in public surveillance. Prespecifying primary reporting pipelines before test unlock, instead of selecting by maximized test ROC-AUC, better matches real deployment conditions-especially given rising SCD prevalence in later years and sparse state coverage in national training years (one to seven states). Concordant locked-test performance across four independent pipelines increases confidence that the benchmark is not an artifact of one language, package, or ensemble rule.

Difficulty deciding was the strongest and most consistent predictor. The BRFSS item asks whether health problems make it hard to make decisions. In survey models, respondents answering “No” (DECIDE = 2) had substantially lower odds of SCD than those answering “Yes” (DECIDE = 1; aOR approximately 0.13; Table 6), so the “Yes” response marks higher SCD risk. This should not be read as a clinical diagnosis of executive dysfunction, and the item may partly capture overlapping physical or mental health burden; nonetheless, it carried dominant signal in ranking, explanation, regression, and graph analyses.

Depression was also linked to SCD in both machine learning and regression analyses, consistent with prior evidence of a strong depression-SCD association [29]. Depression may overlap with SCD symptoms or identify a group at higher long-term cognitive risk. Poorer self-rated general health was another stable correlate, in line with studies linking perceived health to SCD and cognitive health reporting [30–32]. Together with functional limitation items, these findings point to a compact set of already-collected BRFSS measures that track SCD reporting at the population level.

The intended use case is population screening and surveillance, not clinic-based dementia diagnosis. ROC-AUC near 0.76-0.77 indicates useful group-level risk ranking, but individual classification remains imperfect. At validation-tuned Youden thresholds, sensitivity was about 0.58 nationally and 0.51 in New York, with higher specificity (0.82 - 0.87) in both cohorts (Table 5). Operating points can be shifted toward higher sensitivity when case capture is prioritized, including precision-recall-oriented thresholds under class imbalance. Because people with SCD often score normally on brief cognitive screens, models based on broader BRFSS health and function items offer a complementary surveillance signal rather than a substitute for clinical assessment.

Beyond discrimination, the primary pipelines produced usable probability scores. On the locked test set, predicted probabilities agreed reasonably with observed SCD rates (Brier 0.118 nationally and 0.112 in New York; Figure 6A-B), and scores were higher for SCD-positive than SCD- negative respondents (Figure 6C-D). The same score separation persisted across survey years in validation and test data (Figure 7), even as unweighted SCD prevalence rose later. For surveillance dashboards and year-to-year monitoring, calibrated risk scores may therefore be more informative than hard Yes/No labels alone. Thresholds and calibration should still be refreshed as new BRFSS cycles accrue and as Cognitive Decline module geography continues to expand.

Interpretability analyses reinforced the same hierarchy of signals. Difficulty deciding dominated permutation importance and SHAP summaries for the primary pipelines (Figure 8A-B), showed the largest multivariable survey association (Table 6; Figure 8C-D), and sat nearest SCD in both data-only and ML-integrated knowledge graphs (Figure 9; Supplementary Figure S6). Concordance across ranking, local explanation, regression, and network views strengthens the scientific case for a stable BRFSS SCD signature centered on decision difficulty, depression, and perceived health, while remaining explicitly associational rather than causal.

Practically, the open Python/R workflow is meant to be reused: agencies or research groups can refit on local BRFSS extracts, retune thresholds for program goals, and update models when new survey years arrive. The main scientific contribution is therefore not a single classifier, but a temporally disciplined, multi-implementation benchmark showing that future-year SCD risk ranking is achievable from routine surveillance items with convergent interpretability.

### 4.1. Limitations

This study has several limitations. First, BRFSS is cross-sectional and SCD is self-reported, so associations cannot establish causality. We also did not evaluate progression to mild cognitive impairment or dementia; the models therefore rank contemporary SCD reporting rather than future clinical conversion. Second, the machine learning pipelines did not incorporate BRFSS survey weights. National GLMs were unweighted because training-year state coverage was sparse, whereas New York GLMs were design-weighted; weighted and unweighted estimates should not be treated as interchangeable. Third, national training years included only one to seven states, whereas later years had much broader fielding, creating geographic domain shift in addition to rising SCD prevalence. Even with acceptable locked-test calibration, absolute probabilities can drift under such shifts, so periodic recalibration and threshold retuning remain advisable. Fourth, ComBat-lite year offsets were estimated on training years only; validation and test rows received train-fitted imputation without year-mean subtraction for unseen cycles. This choice preserves temporal transport but cannot remove batch structure unique to later survey years. Fifth, locked-test sensitivity at Youden thresholds was only moderate (about 0.58 nationally and 0.51 in New York; Table 5), so hard classification is imperfect for case finding unless operating points are deliberately shifted toward higher sensitivity. Soft-voting and single-model pipelines were nearly tied on ROC-AUC; we therefore emphasize robustness across implementations rather than small rank-order differences. Finally, knowledge-graph displays and survey regressions are associational complements to the prediction pipelines-not causal maps and not direct repeats of SHAP or permutation rankings. Future work should incorporate survey-weighted learning where design variables allow, evaluate transport to newly fielding states, and link BRFSS-based SCD risk scores to longitudinal cognitive outcomes when such linkages become available.

## 5. Conclusions

We developed and temporally validated an open multi-language machine learning benchmark for self-reported subjective cognitive decline (SCD) in large BRFSS cohorts from the United States and New York. Using chronological training, validation-only algorithm selection and calibration, and a locked 2023-2024 test evaluation across four Python/R pipelines, the prespecified primary models achieved consistent future-year risk ranking (locked-test ROC-AUC approximately 0.76- 0.77), with usable calibrated probabilities and persistent score separation across survey years. Interpretability analyses converged on difficulty deciding, depression, and self-rated general health as central signals. The intended use is population surveillance and hypothesis generation-not clinical dementia diagnosis. The released workflow offers a reproducible template for agencies and researchers to refit, recalibrate, and update SCD risk models as new BRFSS cycles accrue.

## Data Availability

All data produced are available online at: https://www.cdc.gov/brfss/index.html
BRFSS datasets are public via CDC. BRFSS is a public, de-identified surveillance dataset; institutional review board approval was not required for secondary analysis.

## Acknowledgements

We thank the CDC Behavioral Risk Factor Surveillance System program and participating state health departments for making the public survey data available. We also thank the Research Capacity Core, CUNY School of Medicine, for providing computational infrastructure and support for model development and the machine learning/AI analyses in this study.

## Data and code availability

BRFSS datasets are public via CDC. BRFSS is a public, de-identified surveillance dataset; institutional review board approval was not required for secondary analysis. Code, processed cohort files, pipeline and analysis scripts are available at https://github.com/truong128/brfss-scd-ml-benchmark.

## Author contributions

T.T.N. and T.D.N. designed the study, performed the data analyses, and drafted the manuscript.

T.T.N. and T.D.N. wrote and revised the manuscript. All authors read and approved the final manuscript.

## Declaration of generative AI and AI-assisted technologies in the writing process

ChatGPT was used for language editing during manuscript preparation. The authors reviewed the final manuscript and take full responsibility for its content.

---

**Supplementary Figure S1.**
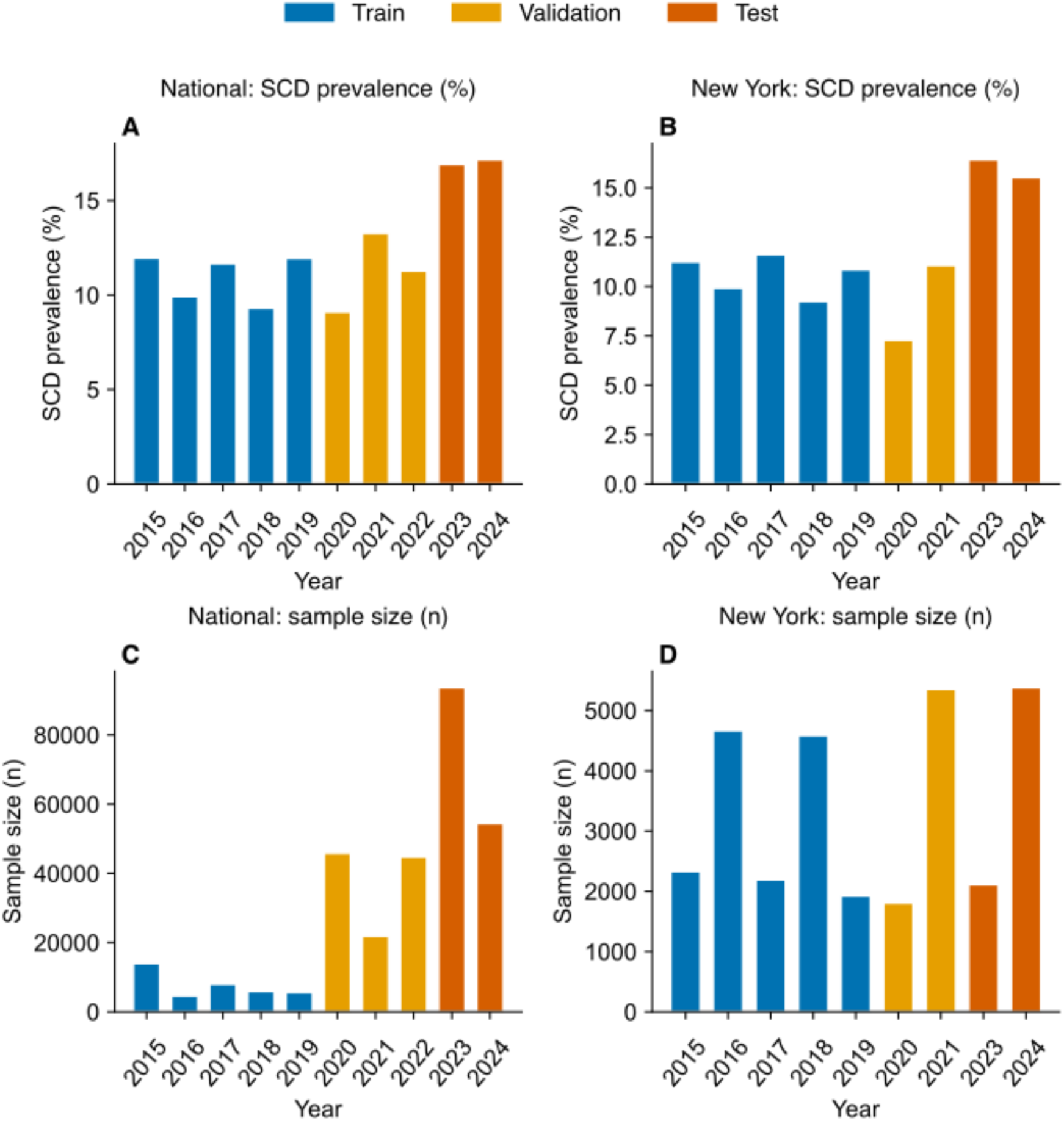
SCD prevalence and sample size by survey year. Unweighted analytic samples from the BRFSS Cognitive Decline module. The national cohort is shown in the left panels (A, C), and the New York cohort is shown in the right panels (B, D). A shared legend denotes temporal splits used in model development: train (2015-2019), validation (2020-2022 nationally; 2020-2021 in New York), and test (2023-2024). (A) National SCD prevalence (%) by survey year. (B) New York SCD prevalence (%) by survey year (2022 data unavailable). (C) National analytic sample size (n) by year. (D) New York analytic sample size (n) by year.

**Supplementary Figure S2.**
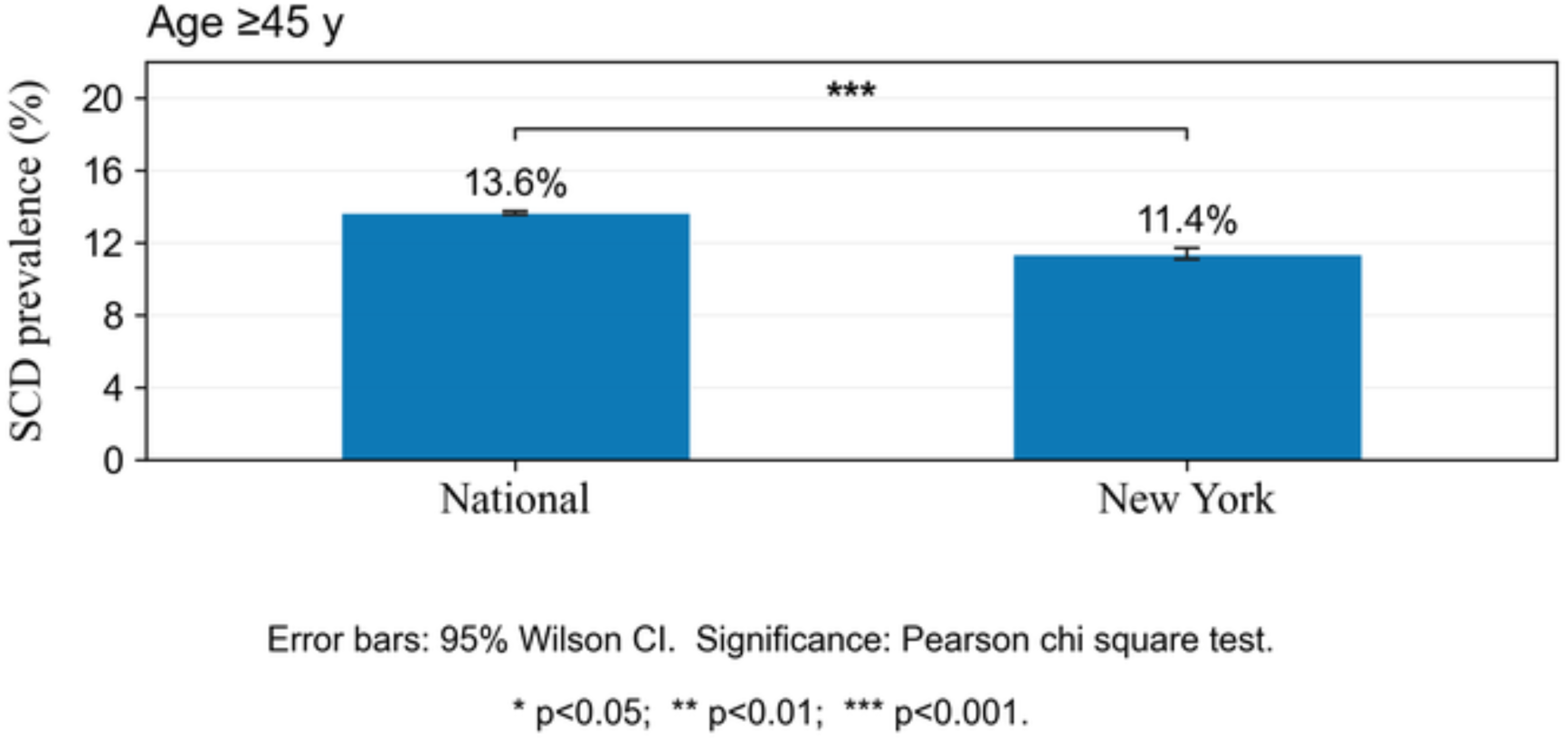
SCD prevalence among adults ≥45 years. Pooled descriptive BRFSS Cognitive Decline module respondents aged ≥45 years across all survey years available for prevalence display (national n = 427,507; New York n = 45,353). These denominators differ from the chronological machine-learning analytic cohorts in Table 1 (national n = 298,944; New York n = 30,366). Side-by-side bars with 95% Wilson confidence interval error bars show unweighted SCD prevalence of 13.6% nationally (58,345 of 427,507) and 11.4% in New York (5,167 of 45,353; P < 0.001). Error bars denote 95% Wilson CIs; the bracket denotes a Pearson χ² test on a 2 × 2 table.

**Supplementary Figure S3.**
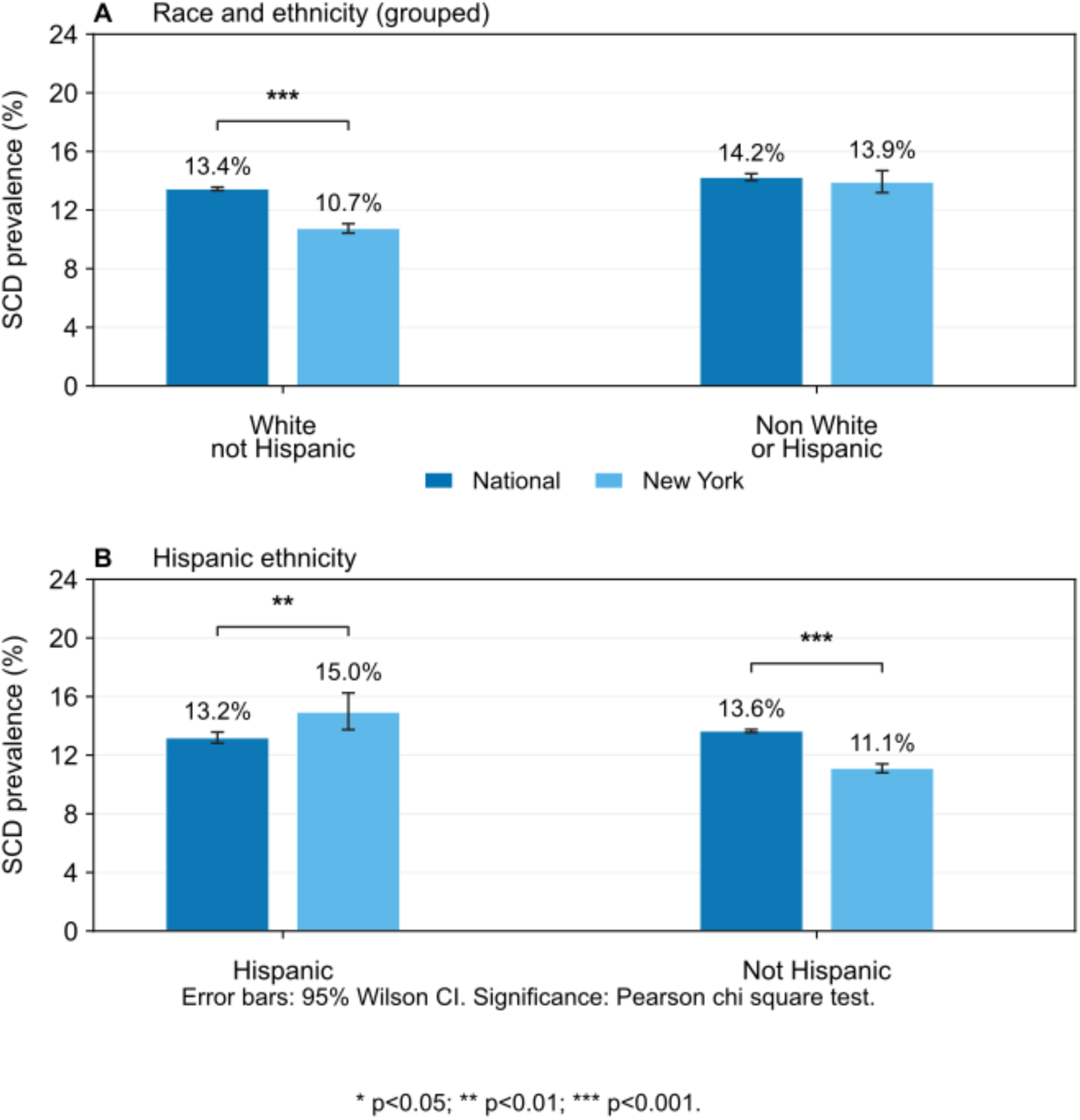
SCD prevalence by race and ethnicity. Pooled BRFSS Cognitive Decline module analytic sample. National estimates are shown in blue and New York estimates in light blue; significance brackets indicate comparisons between cohorts within each stratum. (A) Race and ethnicity (grouped): White non-Hispanic adults had an SCD prevalence of 13.4% nationally compared with 10.7% in New York (P < 0.001), whereas non-White or Hispanic groups showed no significant difference between cohorts (14.2% vs. 13.9%). (B) Hispanic ethnicity: SCD prevalence among Hispanic adults was higher in New York than nationally (15.0% vs. 13.2%; P < 0.01); among non-Hispanic adults, prevalence was lower in New York than nationally (11.1% vs. 13.6%; P < 0.001).

**Supplementary Figure S4.**
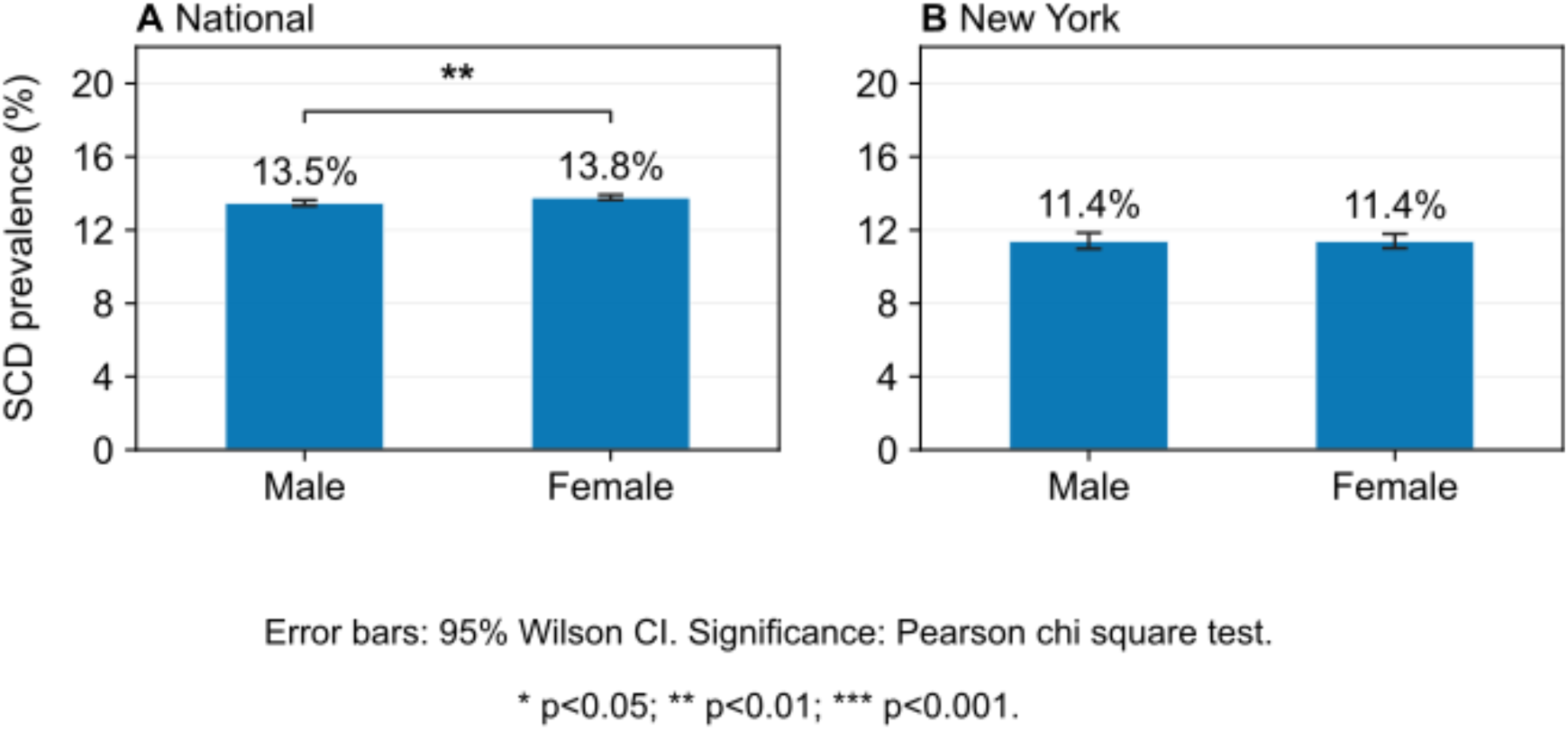
SCD prevalence by sex. Pooled BRFSS Cognitive Decline module analytic sample. (A) National cohort: SCD prevalence was 13.5% among males and 13.8% among females (P < 0.01). (B) New York cohort: SCD prevalence was 11.4% for both males and females (not significant). Error bars denote 95% Wilson CIs; brackets denote Pearson χ² tests.

**Supplementary Figure S5.**
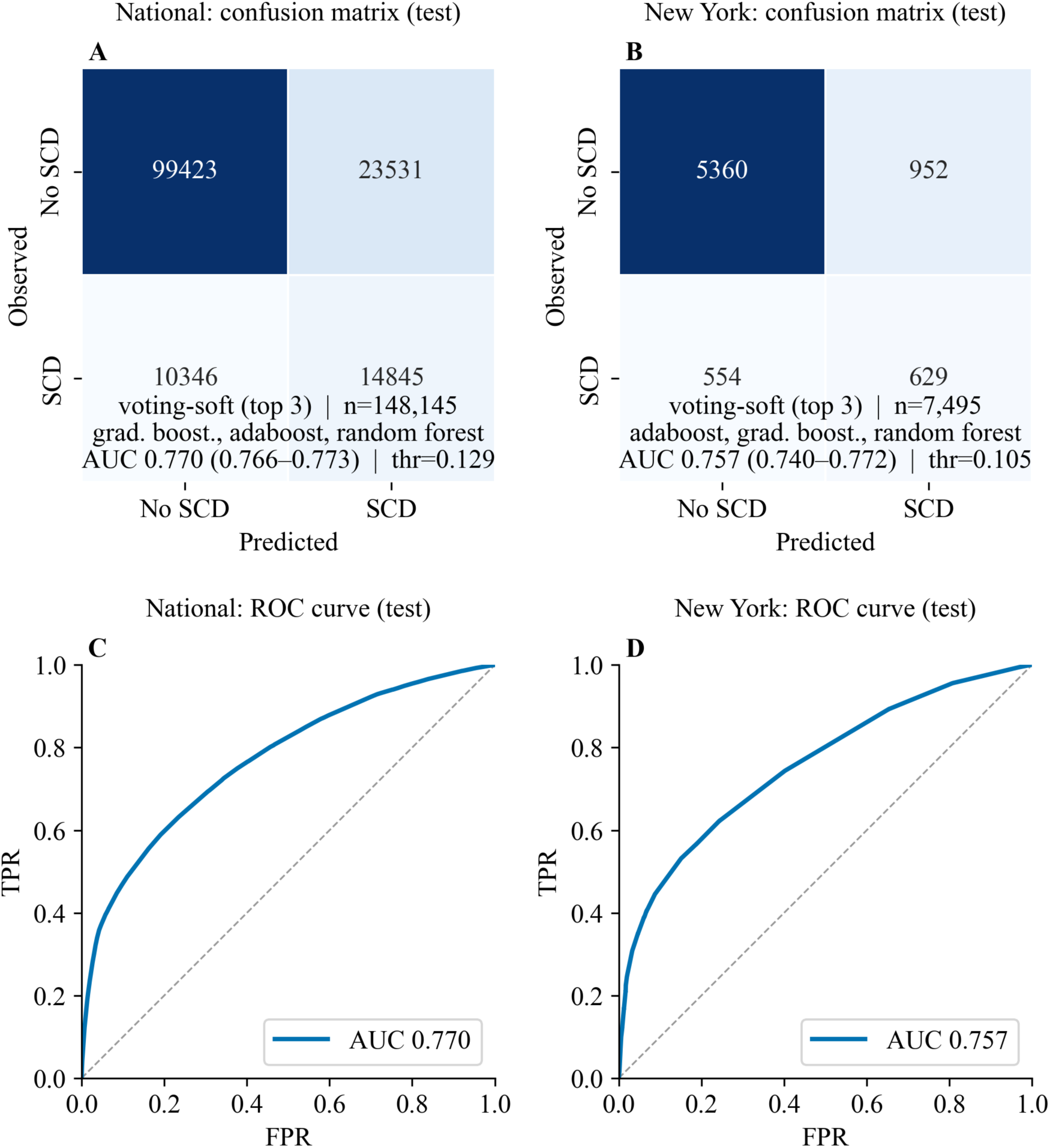
Locked test-set discrimination of the Python soft-voting ensemble (top-3 models, 2023-2024). Companion to Figure 5 using soft-voting of the top three validation- ranked classifiers for both cohorts. National results are shown on the left (A, C) and New York results on the right (B, D). (A, B) Confusion matrices at the validation-tuned probability threshold, with in-panel footnotes for ensemble members, sample size, test ROC-AUC (95% CI), and threshold. (C, D) ROC curves from the same locked test predictions. Soft-voting ROC-AUC values are also summarized in Table 4.

**Supplementary Figure S6.**
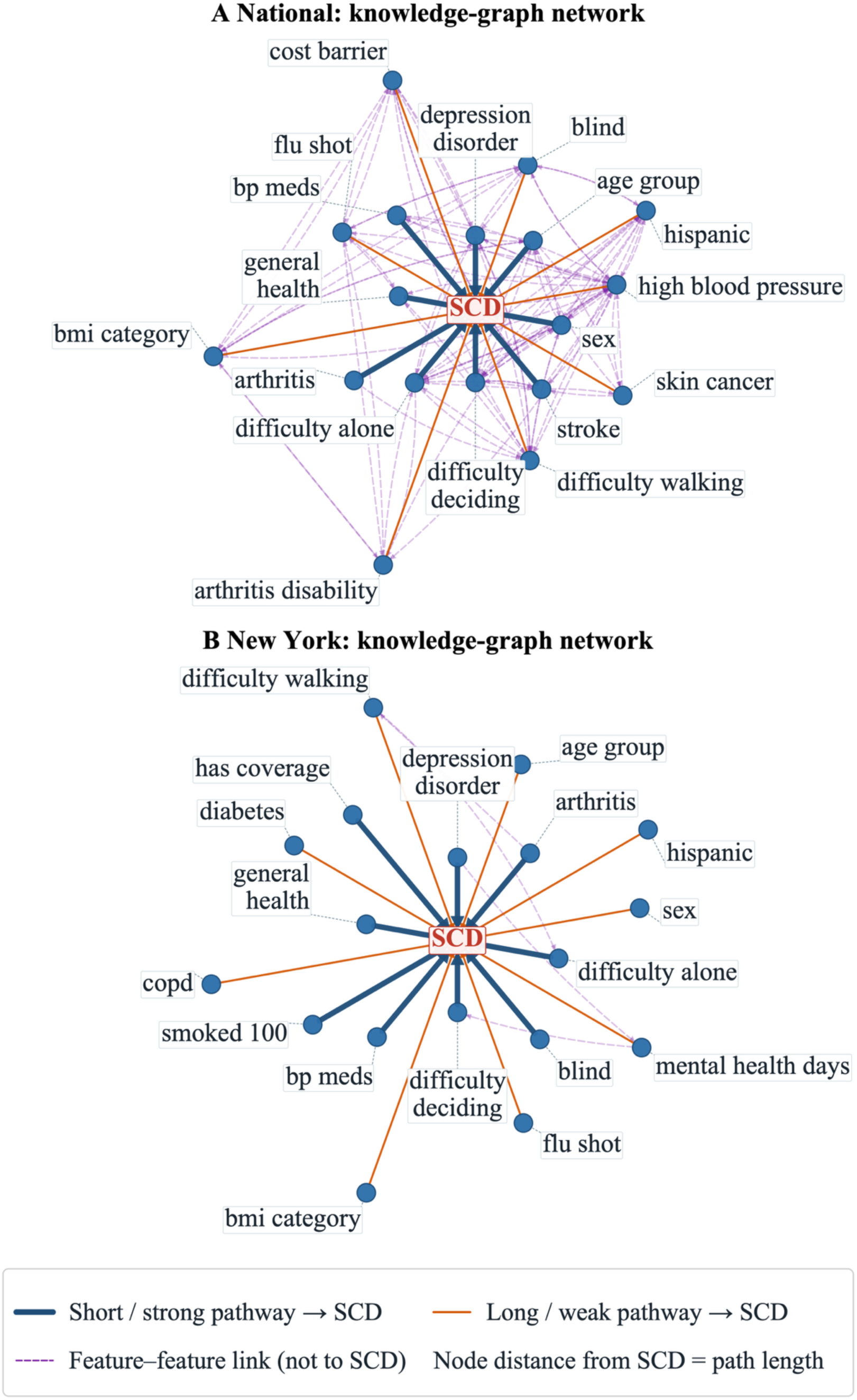
ML-integrated knowledge-graph associations with subjective cognitive decline (SCD). National (A) and New York (B). Graphs integrate training-split GLM associations (national: unweighted; New York: design-weighted; same sources as Table 6), validation-set permutation importance from the Python single-model pipeline, pairwise partial correlations, and domain ontology priors (associational relationships only, not causal relationships). The central red node represents SCD; peripheral nodes are the 18 LASSO-selected predictors with the highest composite association scores. Radial node distance indicates shortest- path graph distance to SCD. Dark blue solid lines, shorter/stronger associational links to SCD; orange solid lines, longer/weaker links; purple dashed lines, feature-feature associations that do not directly connect to SCD.

